# Cross-disciplinary rapid scoping review of structural racial and caste discrimination associated with population health disparities in the 21^st^ Century

**DOI:** 10.1101/2024.01.16.24301383

**Authors:** Drona P. Rasali, Brendan M. Woodruff, Fatima A. Alzyoud, Daniel Kiel, Katharine T. Schaffzin, William D. Osei, Chandra L. Ford, Shanthi Johnson

## Abstract

A cross-disciplinary rapid scoping review was carried out generally following PRIMA-SCR protocol to examine historical racial and caste-based discrimination as structural determinants of health disparities in the 21^st^ century. We selected 48 peer-reviewed full-text articles available from the University of Memphis Libraries database search, focusing on three selected case-study countries-the United States (US), Canada and Nepal. Authors read each article, extracted highlights and tabulated the thematic contents on structural health disparities attributed to racism or casteism. Results linked the historical racism/casteism to health disparities occurring in Blacks and African Americans, Native Americans and other ethnic groups in the US, in Indigenous peoples and other visible minorities in Canada, and in Dalits of Nepal, a population racialized by caste, grounded on at least four foundational theories explaining structural determinants of health disparities. The evidence from the literature indicates that genetic variations and biological differences (e.g., disease occurrence) occur within and between races/castes for various reasons (e.g., random gene mutations, geographic isolation, and endogamy). However, historical races/castes as socio-cultural constructs have no inherently exclusive basis of biological differences. Disregarding genetic discrimination based on pseudo-scientific theories, genetic testing is a valuable scientific means to achieve better health of the populations. Epigenetic changes (e.g., weathering – early aging of racialized women) due to DNA methylation of genes among racialized populations are markers of intergenerational trauma due to racial/caste discrimination. Likewise, chronic stresses resulting from intergenerational racial/caste discrimination cause ‘allostatic load,’ characterized by an imbalance of neuronal and hormonal dysfunction, leading to occurrences of chronic diseases (e.g., hypertension, diabetes, mental health) at disproportionate rates among racialized populations. Major areas identified for reparative policy changes and interventions for eliminating health impacts of racism/casteism include health disparity research, organizational structures, programs and processes, racial justice in population health, cultural trauma, equitable healthcare system, and genetic discrimination.

**Highlights:**

- Research on the relationship between structural racism and population-level health outcomes and on the health impacts of policy changes and interventions has largely overlooked caste, which is a system of racialization that pre-dates US racial categories.
- A cross-disciplinary global ‘caste’ approach is adopted to examine various forms of historical decent-based racial and caste discrimination in three case-study countries.
- Major theories and praxis explaining research and experiences of structural racial and caste discrimination impacting health disparities are consolidated to synthesize a unified body of knowledge for the 21^st^ century.
- While genetic variations occur naturally, they do not inherently contribute to the social constructs of race or caste.
- Reparative policy changes and interventions are necessary to eliminate deeply entrenched structural health disparities rooted in racism and casteism.

## 1. Background

Health disparities within and between populations, across geographic regions, demographic groups such as age and sex, and due to socio-economic factors such as income, education, employment, complex indices of material and social deprivation, and environmental factors have been well documented in peer-reviewed literature (Zhang et al., 2015; Zandy et al., 2019; GBD US, 2022; Relova et al., 2022) and gray literature (CDC, 2011; Rasali et al., 2016; Rasali et al., 2019). In the United States (US), Global Burden of Disease and US Health Disparities Researchers have found that life expectancy disparities between racial-ethnic groups are widespread and persistent and have stressed that analysis of local-level data is vital to address the root causes of poor health and premature death among disadvantaged groups and to eliminate health disparities (GBD US, 2022). However, these reports merely address “risk-factor or individual social determinants of health (SDoH)” based on epidemiological data that show plainly the associated differences between advantaged and disadvantaged groups but do not provide concrete evidence of the root cause of these racial health inequities, which are unfair, avoidable, and unjust (Melton-Fant, 2022) in the present-day society. Research on the relationship between structural racism and population-level health outcomes and the health impacts of policies and interventions dismantling the former has been limited (Bailey et al., 2017).

Recent anthropological and ancient genome data (Reich, 2018; Narasimhan et al., 2019) indicate that discrimination between groups of people originated during the transition from pastoralism to agriculture in ancient times and has since spread globally. Consequently, various regional race/caste systems have developed over time across the globe, such as *Varnashram-based* caste discrimination and untouchability practiced in South Asia, racism against African Americans and Native Americans in the United States, colonial racism against Indigenous Peoples in Canada, Castes of West Africa, Castas of Latin Americas (Yengde, 2022), and other forms of inherited racial hierarchies still active in East Asia (Kowner and Demel, 2015). Racism, casteism, or all other descent-based social stratification systems have the same core elements: a system of dehumanization, inequality, and condemnation (Yengde, 2022) in which a dominant group overpowers a marginalized group with the sole purpose of holding on to their resources by exercising privilege and power. In the aftermath of the early phase of the COVID-19 pandemic that left excess numbers of African American, Latinx, and Indigenous people among the sick and dying, and the anti-racism protests sparked by murder of George Floyd, Isabel Wilkerson’s book, *Caste: The Origins of Our Discontents* brought widespread attention in the West to the concept of “caste”. Initially, it had been thought that caste was confined to the Indian subcontinent as a phenomenon unrelated to American racism (Bassett, 2021). We surmise that the consideration of historical, deep-rooted, systemic, and structural racism/casteism as the common root cause of racial/caste-based socio-economic and cultural disadvantageousness globally is essential to addressing the health disparities that the whole of humanity currently faces. However, the evidence of the full scope of this phenomenon of descent-based discrimination has not been consolidated. Reconciling the global literature is vital to understand it as a historically created global social construct.

The castes and races, thus hierarchically stratified by humans, and their associated inequalities might have common ancient roots—the transmigration and interactions among ancient cultures that spread to the East and West from Anatolia, the European Steppe, and the Mesopotamian Fertile Crescent (Reich, 2018). Building on the tradition of feudal and religious hierarchies, European colonists developed the concept of racial superiority to justify the needs of European empires as they expanded across the globe (Wallis & Fleras, 2009). European colonists who introduced slavery to the Americas in the early 17^th^ century buttressed the discriminatory caste hierarchy in South Asia through Anglo-Hindu Law based on colonial interpretations of Hindu scriptures and customary law in British India in 1772 and the promulgation of Nepal’s Civil Code in 1854 (Rasali, 2023). In the mid-19^th^ century, books like Robert Knox’s The *Races of Man* (1850) and Arther de Gobineau’s *Essays on Inequality of Human Races* (1853) gave race stratifications a secular, rather than religious explanation through the use of pseudo-science (Wallis & Fleras, 2009), reinforcing racial discrimination that has been ingrained in society ever since.

Western racial discrimination and South Asian caste discrimination, with a common root of origin—the ancient mythical history of Aryan supremacy further advanced during the colonial era from the 17^th^ century onwards (Poliakov, 1974) in both worlds. It is well-documented that Black people and Native Americans are affected by structural racism that have deep historical roots in American society (Bailey et al., 2017). Nepalese anthropologist Dor Bahadur Bista (1991) was convinced that caste discrimination, which persists in Nepal today, is a colonial import since the 18th century. Both, structural racism in America and structural caste discrimination in South Asia, may produce similar health disparities in their populations, though reliable health data on caste discrimination for comparison are limited. Both kinds of discrimination dehumanize and marginalize an oppressed segment of the population, resulting in adverse health outcomes at the societal level through similar modes and mediating factors. In this rapid scoping review, we explore how systems of racial/caste discrimination have led to systemic socio-economic inequalities and population-level health disparities in the 21st century. We used the United States, Canada, and Nepal as country examples for case studies. The United States has a complex history of racism deeply rooted in its past, beginning with slavery in the 17th century, the crude and brutal racism of the antebellum period, and post-reconstruction Jim Crow laws, which segregated Black and White citizens. Despite the achievements of the Civil Rights Movement and the subsequent passage of the Civil Rights Act, racial disparities in health, housing, education, criminal justice, and wealth persist. Likewise, racism against Native Americans has deep historical roots of dispossession, forced assimilation, and cultural suppression (Kirmayer et al., 2014). At the same time, Hispanic and Asian-American populations also face challenges of discrimination, implicit bias, and unequal opportunities (Krieger, 1014).

In contrast to the blatant racism in the United States, Canada has handled race differently, with historically “milder” racism built on a social order of “colorblindness” transitioning to present-day multiculturalism (Wallis and Fleras, 2009). Browne (2017) argues that the ideologies of egalitarianism and multiculturalism in Canada, painting a picture of the state and its institutions as free from racism and other forms of discrimination, perpetuates the assumption that people are treated the same regardless of their diverse backgrounds. Still, structural racism has been extant pervasively in the form of implicit bias and unequal opportunities against Indigenous people and other people of color from various ethnic groups who are known as visible minorities. Nepal is a particular case in the 21st century when the country is transitioning from the crude form of historical hierarchical caste-based discrimination enforced through the Civil Code 1854 law to more recent and progressive constitution and laws banning all forms of discrimination in alignment, in statutory books, with the United Nations Declaration of Human Rights. However, the system of caste discrimination persists in existential realities as a form of structural casteism deeply ingrained in the society, even today, mainly because of weak enforcement of the new laws that severely lack representation from discriminated caste groups in the enforcement bodies. In all three country case studies, we aim to review the current state of health disparities associated with racism or casteism in its structural form in the 21st century. As European powers were colonizing large swaths of the globe, colonial racism allowed wealthy and aristocratic colonizers to consolidate power in the Antebellum United States, Europe, and other colonized lands. However, the American Civil War (1861-1865) changed the course of US history, while colonial powers eventually declined worldwide, especially after World War II, which led to significant global political change as many nations embraced democracy and civil rights. Unfortunately, pseudo-scientific theories advocating falsely a biological basis for racial differences were widely accepted until much later, even by the end of 20^th^ Century. Hence, our review only focuses on scientific literature published in the 21st century, when a scientific consensus around health disparities associated with structural racism disregarding pseudo-scientific theories has consolidated, acknowledging that race and caste discrimination is immoral, despite it still persists in a systemic and structural form.

### Study objectives

The main objectives of this rapid review are the following:

- To collate the literature evidence on the background realities of historical and structural racial and caste discrimination causing geographic, demographic, and socio-economic inequities resulting in population health disparities at the societal level in the context of three case-study countries.
- To draw major themes of societal health disparities based on the theories, emerging concepts, various components, and determinants associated with structural racism and casteism.
- To consolidate a set of recommendations for reparative policy changes to help learn the evidence of existential realities and unlearn and challenge the historical structural racial and caste discrimination systems that lead to population health disparities in the context of the 21^st^ century.

## 2. Methods

This scoping review was completed rapidly due to the short duration available to the first author’s [designated project]. However, we have followed the PRISMA-ScR protocol (Preferred Reporting Items for Systematic Reviews and Meta-Analyses Extension for Scoping Reviews) as a general guideline methodology to synthesize knowledge, using a systematic approach to map evidence on the historical background and contemporary realities of racial and caste discrimination, their impacts on health equity, and identify central themes, theories, sources, and knowledge gaps.

This review aims to draw cross-disciplinary knowledge of systemic and structural racism/casteism and associated health disparities; we primarily used the University of Memphis Libraries’ broad-based multi-disciplinary database (EBSCO database). The first author, as the principal investigator (PI), in consultation with a University Librarian, drew up a set of search terms, focussing on three program interest case-study countries, the United States, Canada and Nepal, as follows: “*SU (health (disparit* or equit* or inequi* or equal* or inequa* or disadvantag*)) AND SU (rac* or caste or ethnic* or cultur* or soci*) AND SU (discriminat* or prejudic* or bias or oppress* or marginal* or depriv*) AND SU (histori* or genetic* or anthropolog*) AND SU (Canada or (United States or US or USA) or India or Nepal or South Asia).”*

Using these search terms, our most recent search, conducted on September 21, 2023, produced 67 full-text English language articles published between 2000 and 2023 that were available through the Libraries’ database. We wanted to limit our study to literature published in the 21^st^ century after pseudo-scientific theories about biological basis to justify racial differences became no longer acceptable in the mainstream knowledge base. The results were then filtered to include only academic and peer-reviewed journal articles, reducing the number of articles to 53. After removing the duplicates, the number of articles came to 32. Again, the PI looked up the PubMed database using the same search terms for articles covering health disparities that are foundational to this review work but could have been missed from the broad database. The relevant articles were added, bringing the total number to 40. Then, PI further looked up the list of references in the selected articles to identify any articles foundational to structural racism or casteism, adding 11 articles to the collection, and three manuscripts were excluded for lack of relevance, bringing the final number of selected articles to 48. The flowchart diagram of our search strategy is presented in Figure 1.

**Figure 1.**
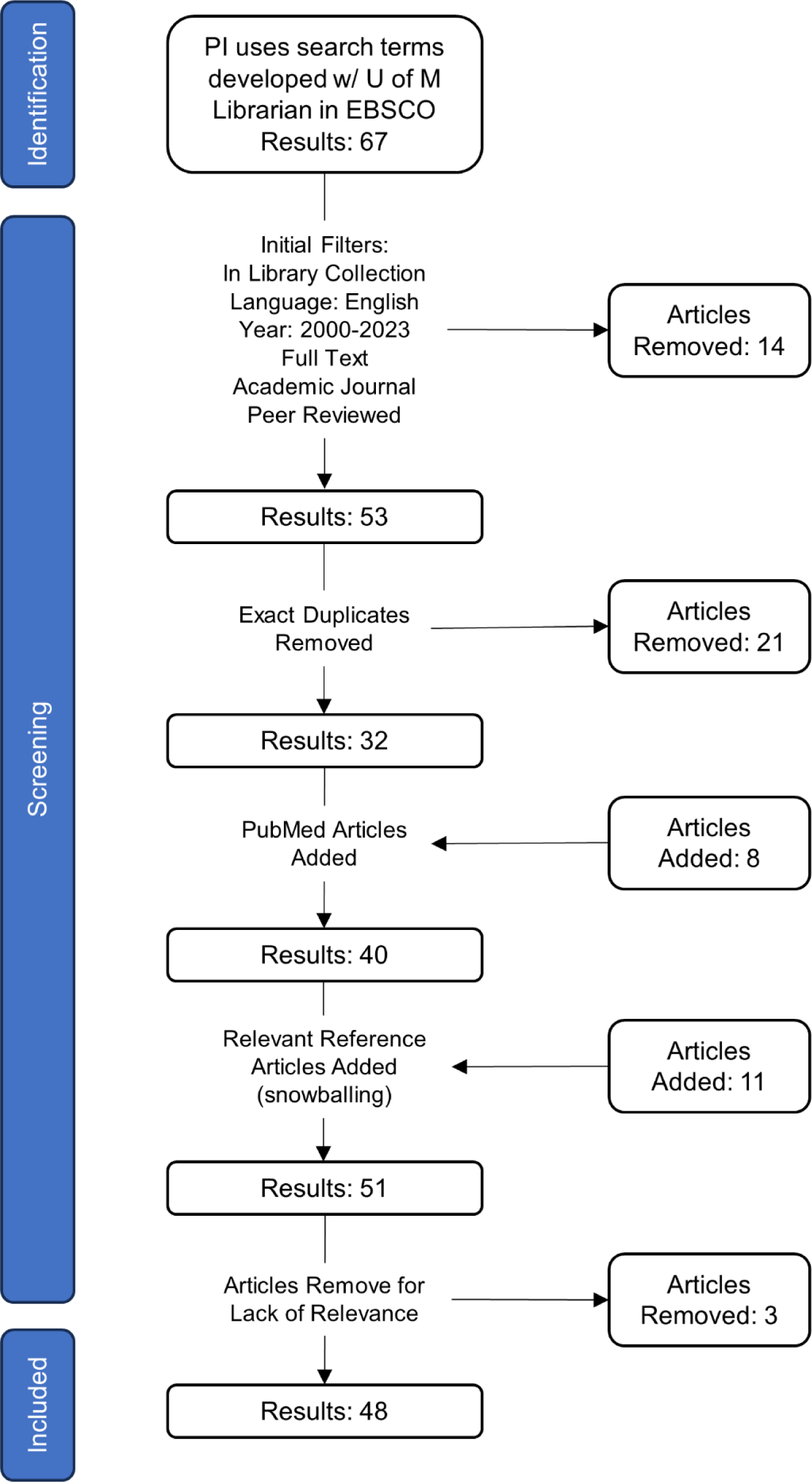
Flowchart of literature search strategy.

The PI read all the selected articles and highlighted the relevant parts in the text for extraction. The Research Assistant (RA) extracted all the highlighted texts from the selected papers into separate documents using an Adobe Acrobat plug-in. RA then created an Excel Worksheet to list all the selected articles, with their title, authors, abstracts, and extracts from the highlighted parts of articles. The RA compiled the thematic items collaboratively with the PI to summarize the excerpts. Various themes were drawn manually and intuitively from the extracts that were categorized and separated as thematic items into worksheet columns and directly from the articles that will be divided into the Worksheet columns. RA put together the search flowchart and tables. The PI wrote the text for the first draft, and RA compiled the tables and figures from the extracts. The first draft was saved into a shared Google Doc folder for everyone among authors to access and provide revision inputs and comments for improvement. All documents (search strategy, selected articles, protocol, worksheets, manuscript drafts) collected were saved in the shared Google Doc folder(s) that were accessible to all the authors.

Other authors contributed to the writing of the manuscript, at the stage of editing with their comments and direct revisions, for the preparation of the second draft of the manuscript. All authors accessed and read the final version of the manuscript that was shared in a Google drive folder to review draft, offered insightful inputs for revision, and agreed to be co-authors for publication in an international, cross-disciplinary journal.

## 3. Results and Discussion

The final analytic sample included 48 articles. Table 1 presents in the thematic characteristics extracted from them. Twenty-one articles dealt with health disparities across various forms of racism and casteism. Nine articles discussed health disparities impacting Black African Americans; two discussed health disparities impacting Indigenous peoples; four discussed racism-related health disparities impacting other ethnic groups; four discussed caste-based health disparities and the link between casteism, transnational casteism, and racism; and two discussed transnational casteism. Twelve articles dealt with the theoretical underpinnings of research into race and caste-based health disparities. Twelve articles dealt with genetics–with ten articles discussing genetic variations and two discussing genetic testing. Three articles dealt with epigenetic changes. Thirteen articles dealt with psychosocial stresses, allostatic load, and telomere shortening. Fifteen articles dealt with social determinants of health and structural determinants of health disparities. Finally, eighteen articles dealt with reparative policy considerations. Thus, structural racism or casteism has been recognized as a determinant of health disparities in at least eight articles reviewed (Bloche, 2004; Gee & Ford, 2011; Browne et al., 2016; Bailey et al., 2017; Williams et al., 2019; Martinez et al., 2021; Diaz et al., 2023; Sweeting et al., 2023). Other articles reviewed specifically included six reports on genetic variations, genetic testing, and epigenetics, seven reports on racial discrimination in Indigenous peoples, and three articles on intergenerational trauma. We summarized below some of the major themes that emerged from the review synthesis in the context of this research on structural racism/casteism as the root or structural determinant of health disparities in the 21^st^ century, with unique case-study references to the United States, Canada and South Asia, particularly Nepal.

**Table 1.**
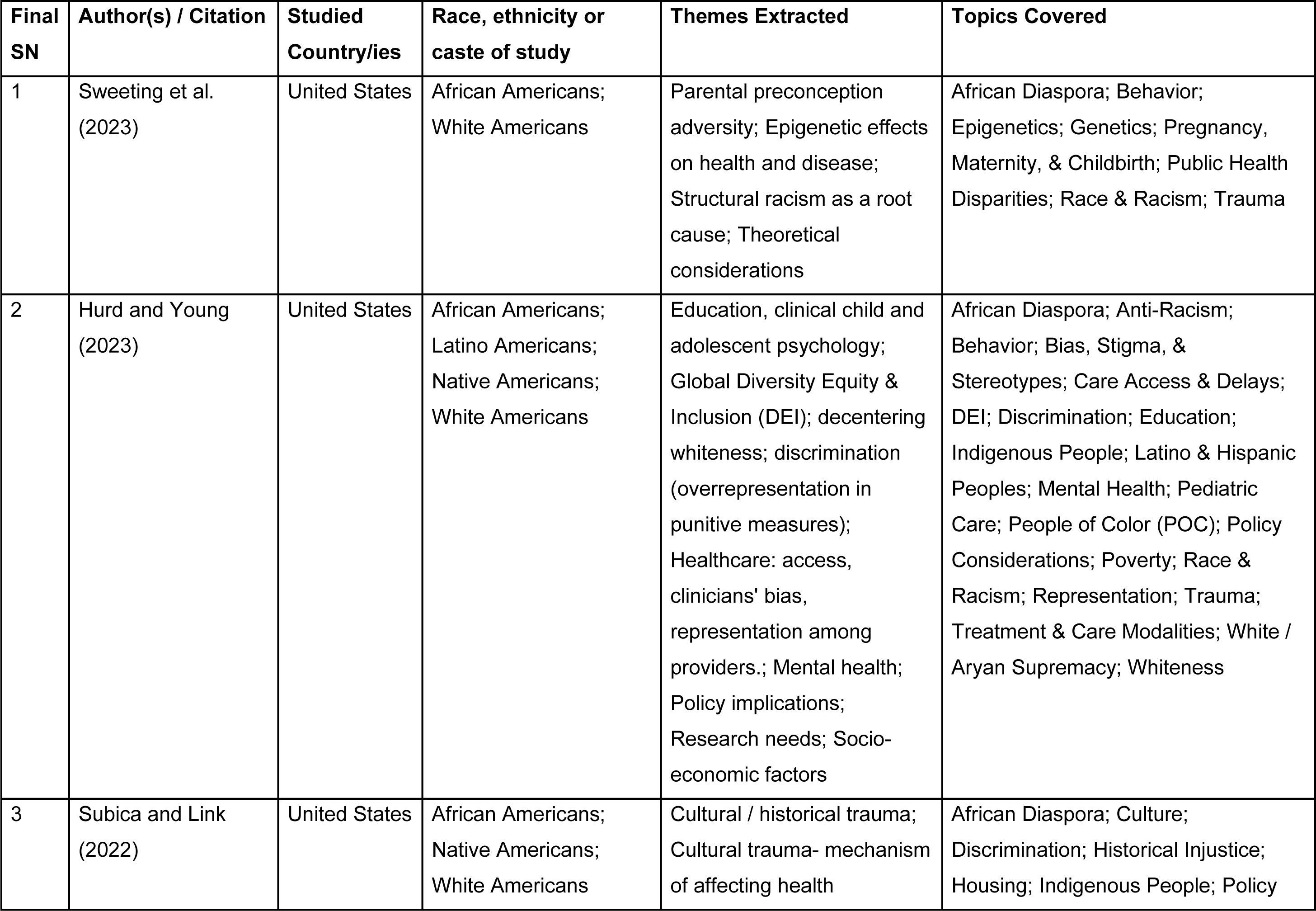
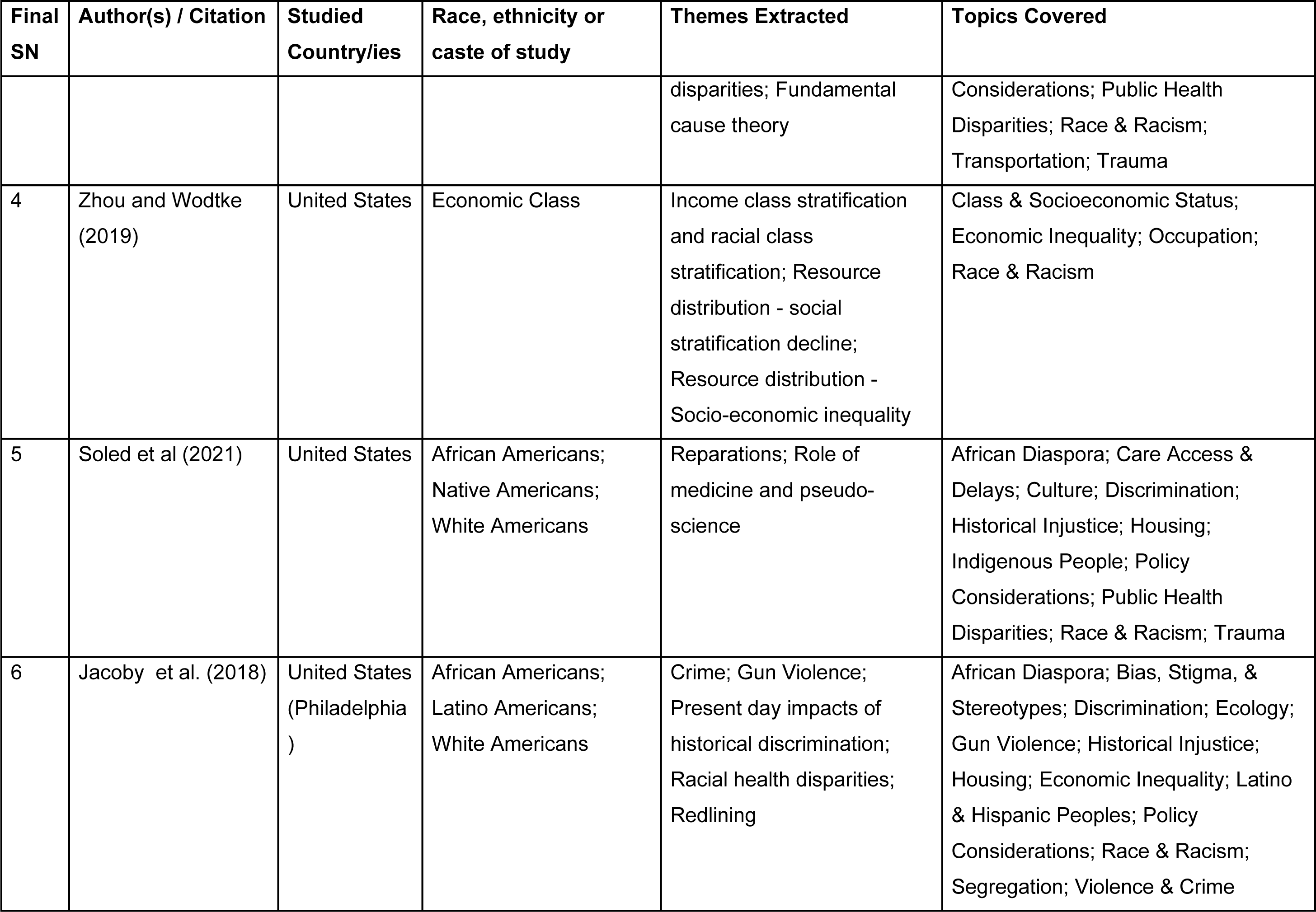
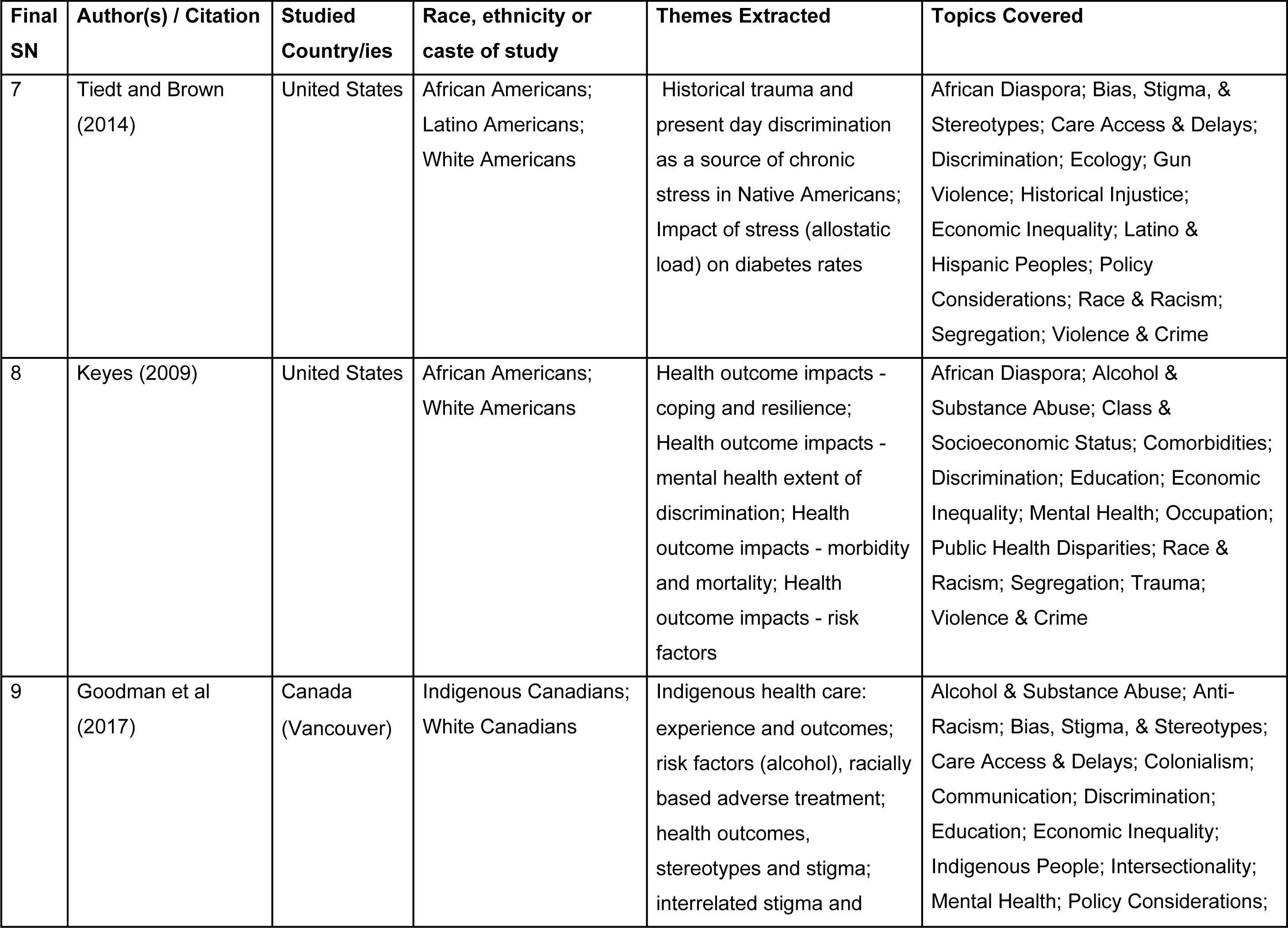
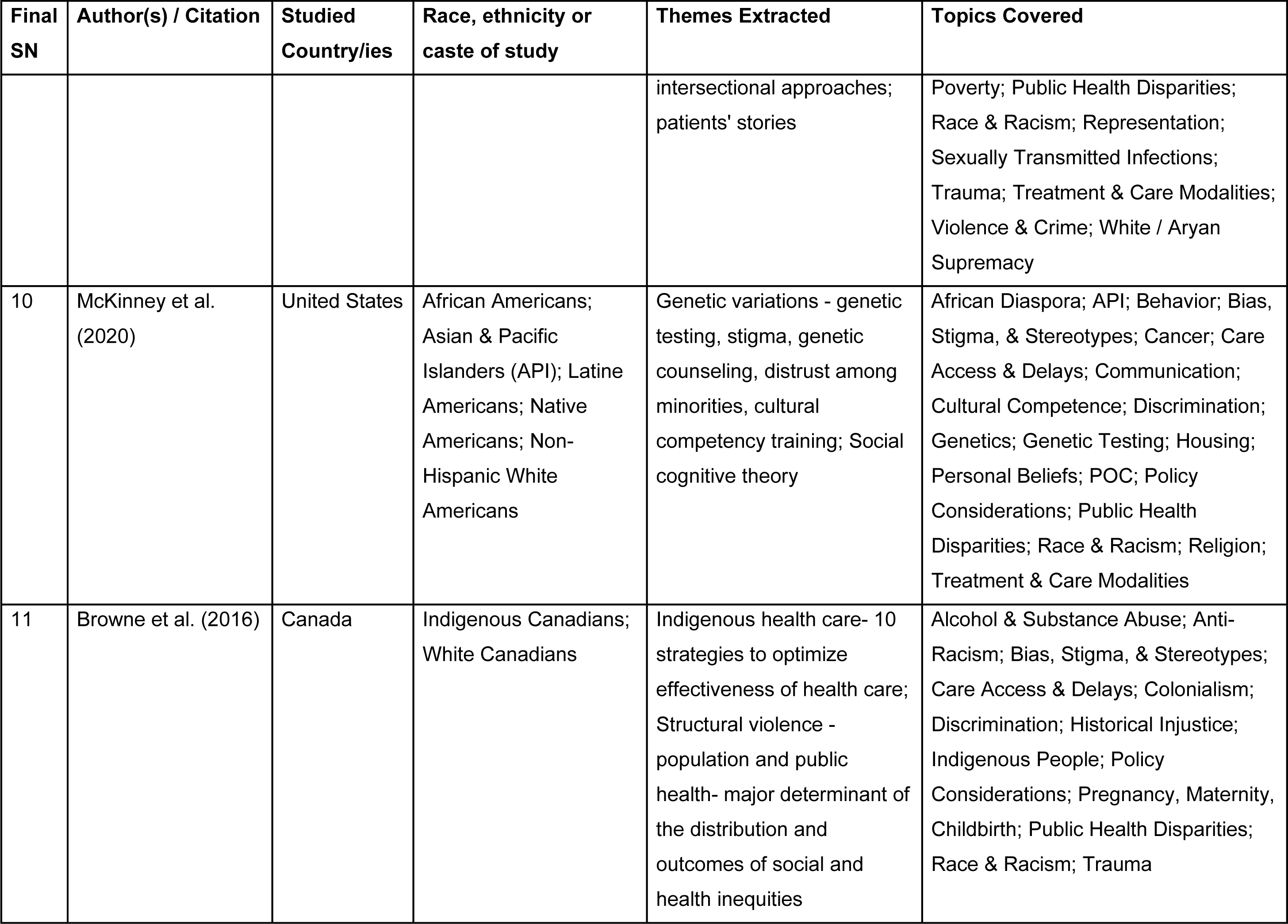
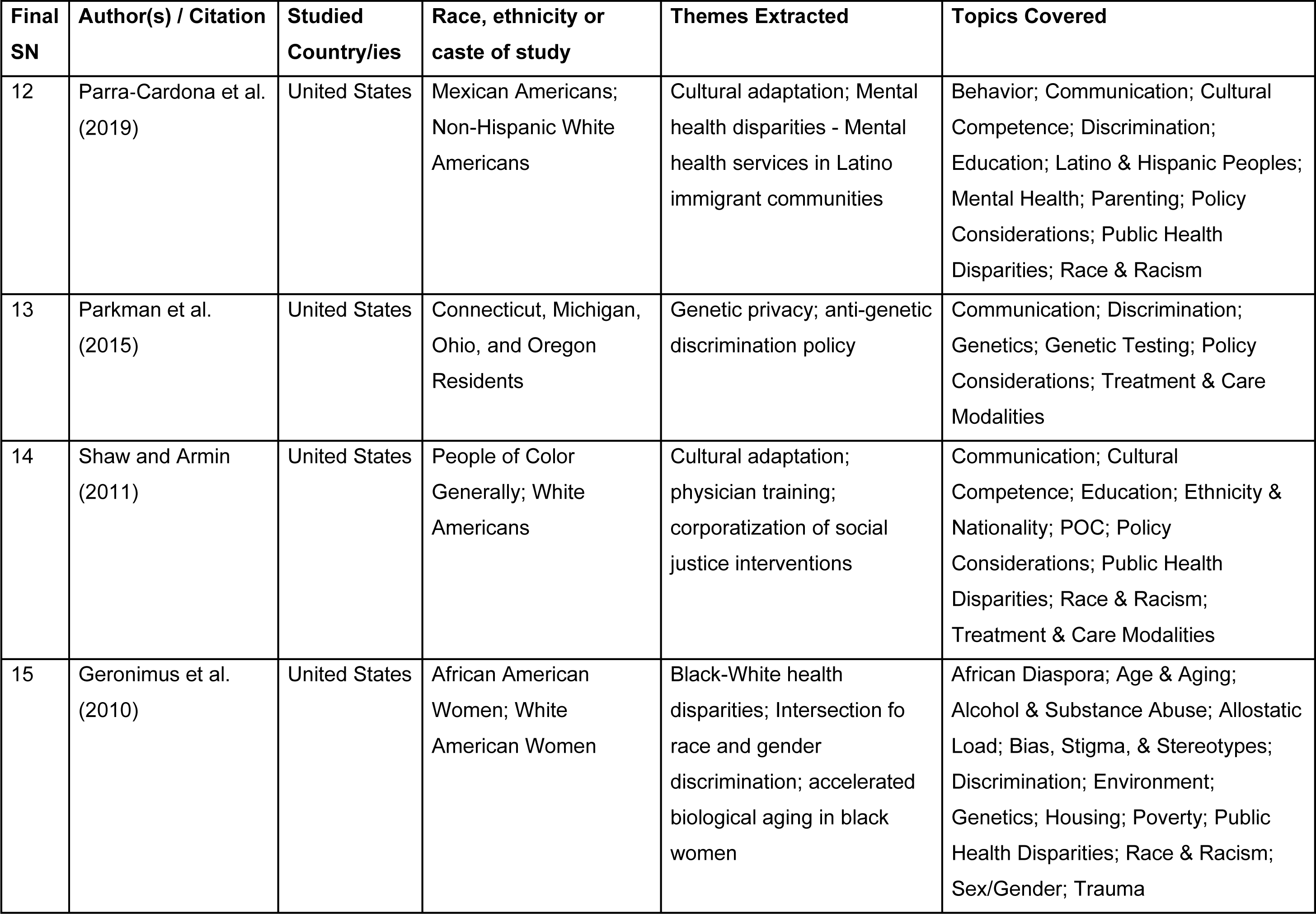
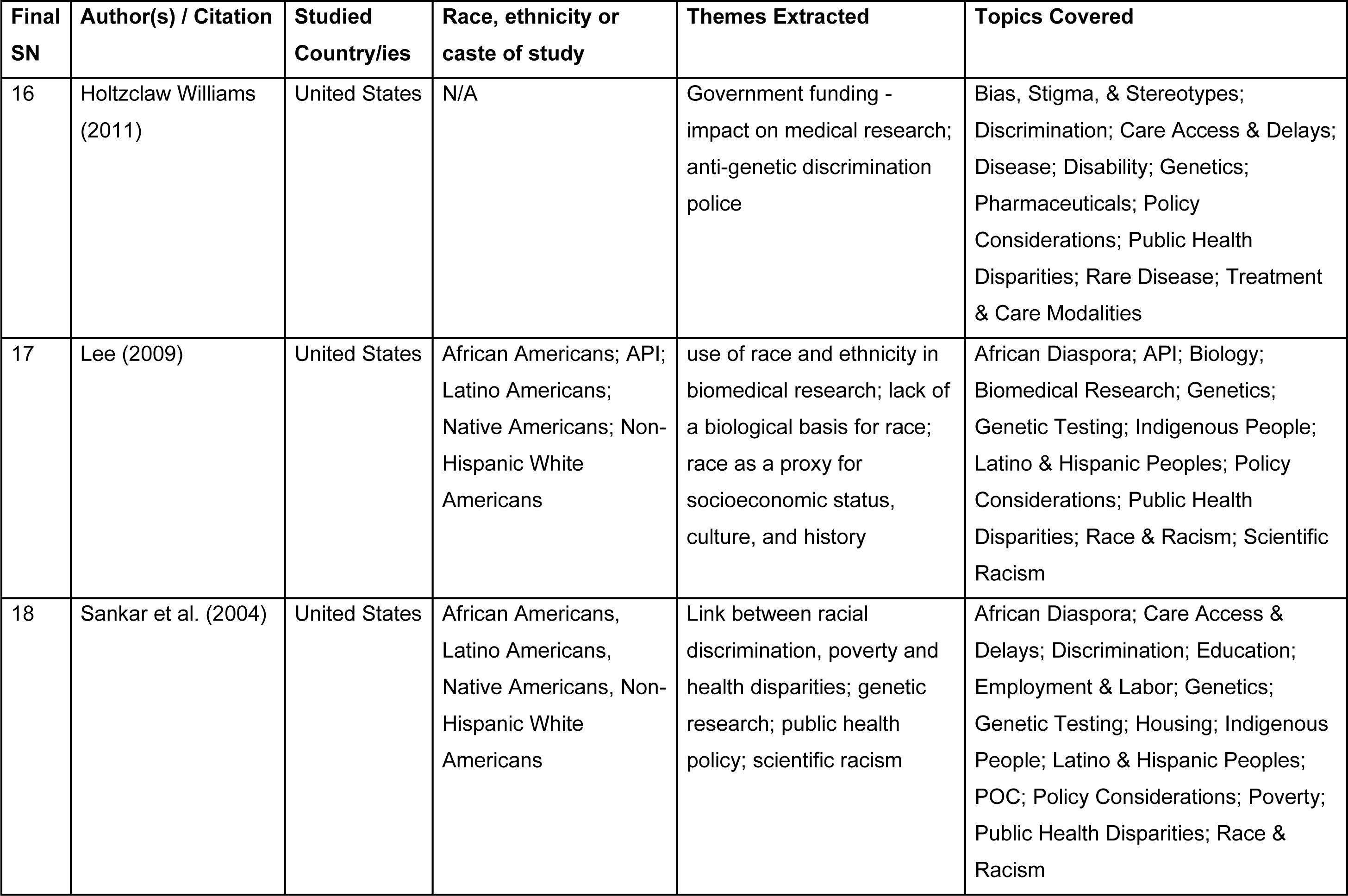
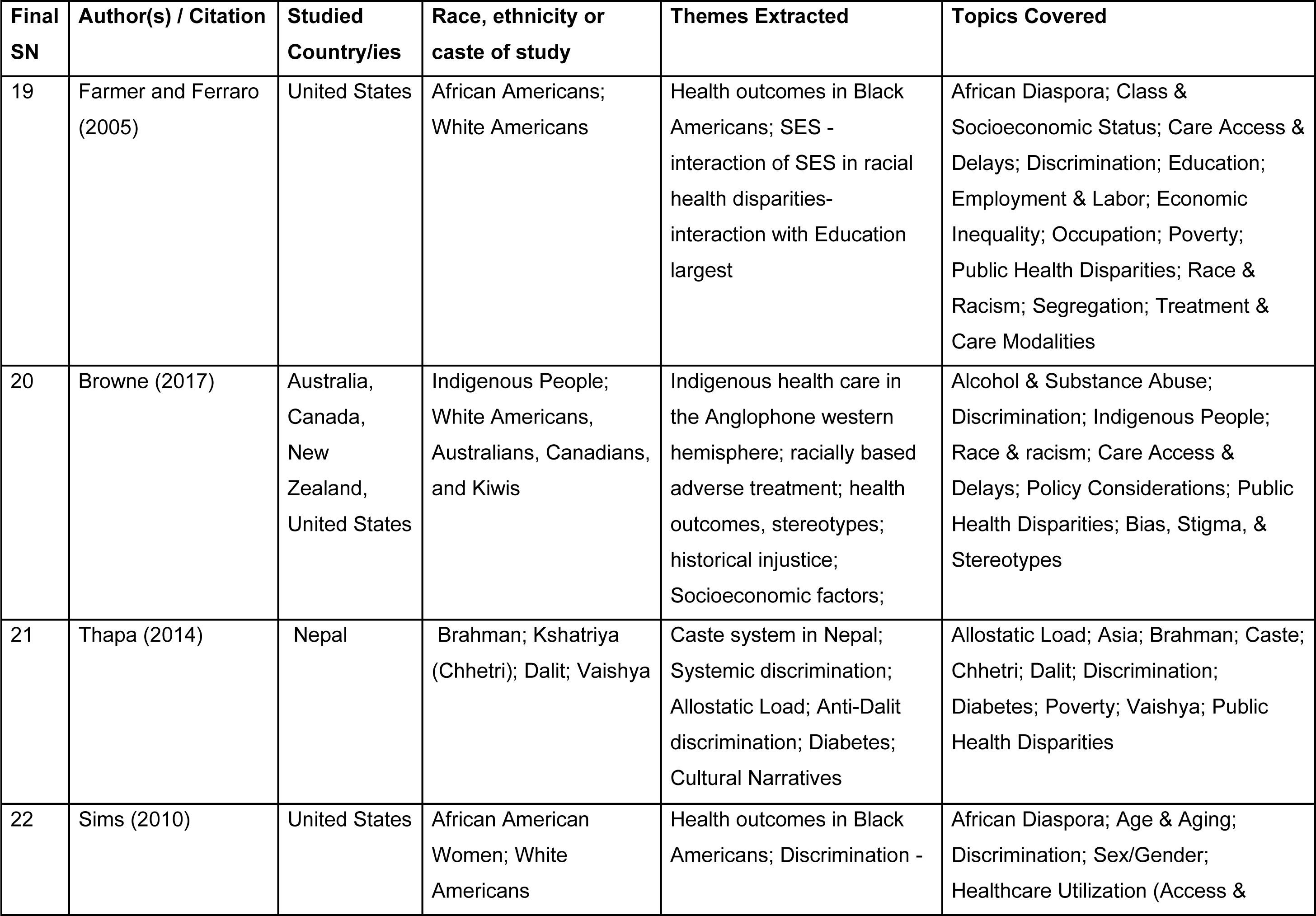
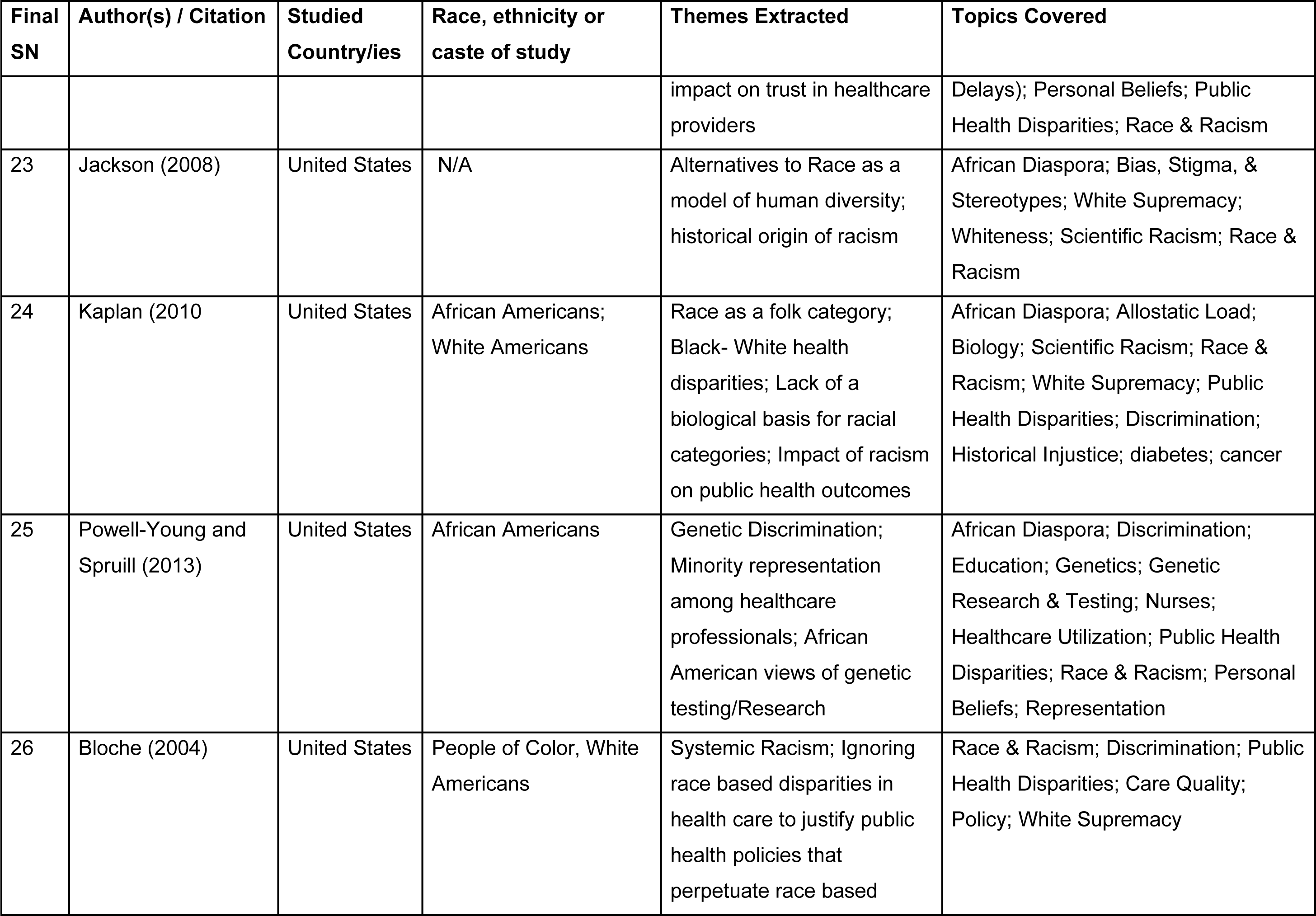
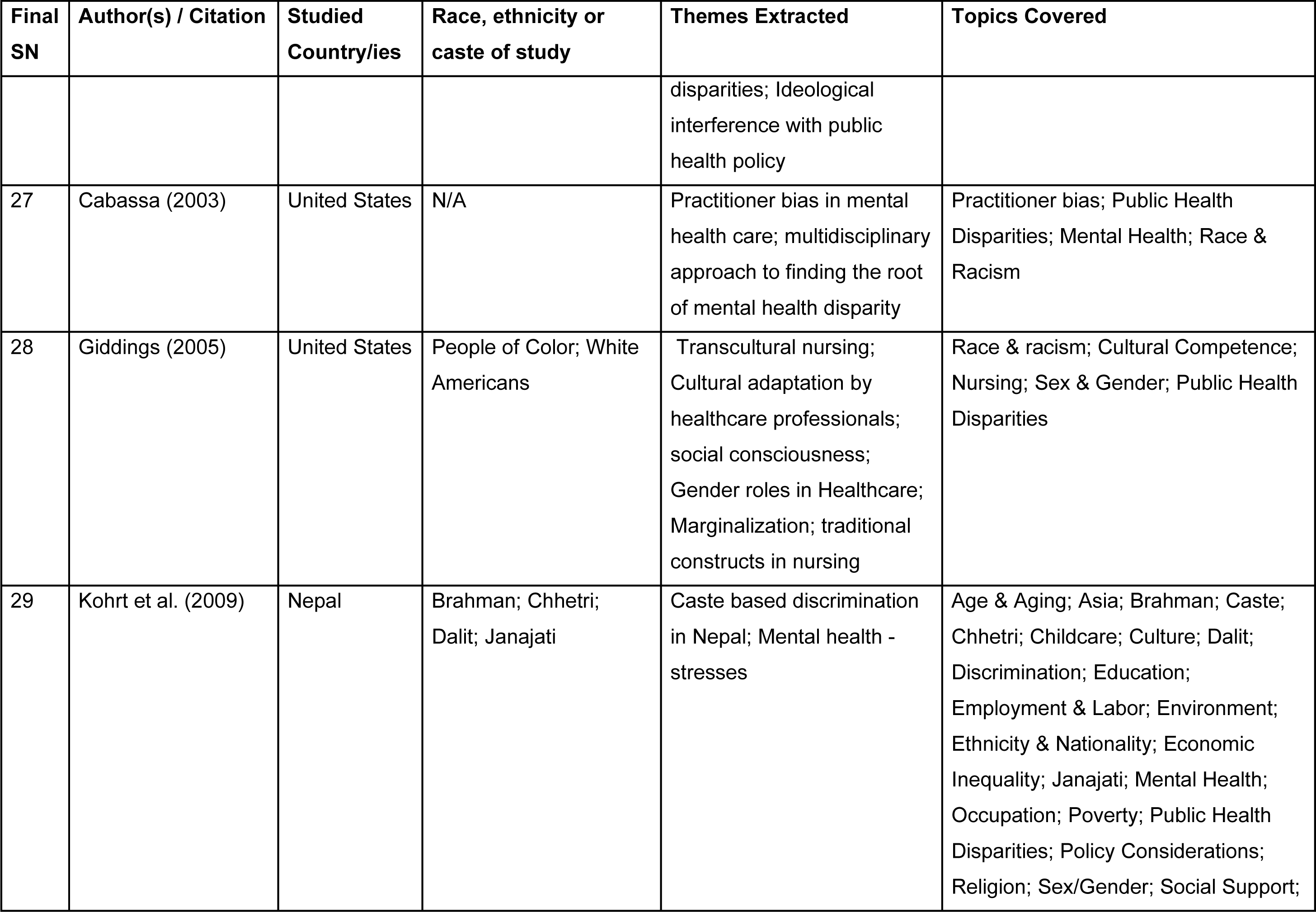
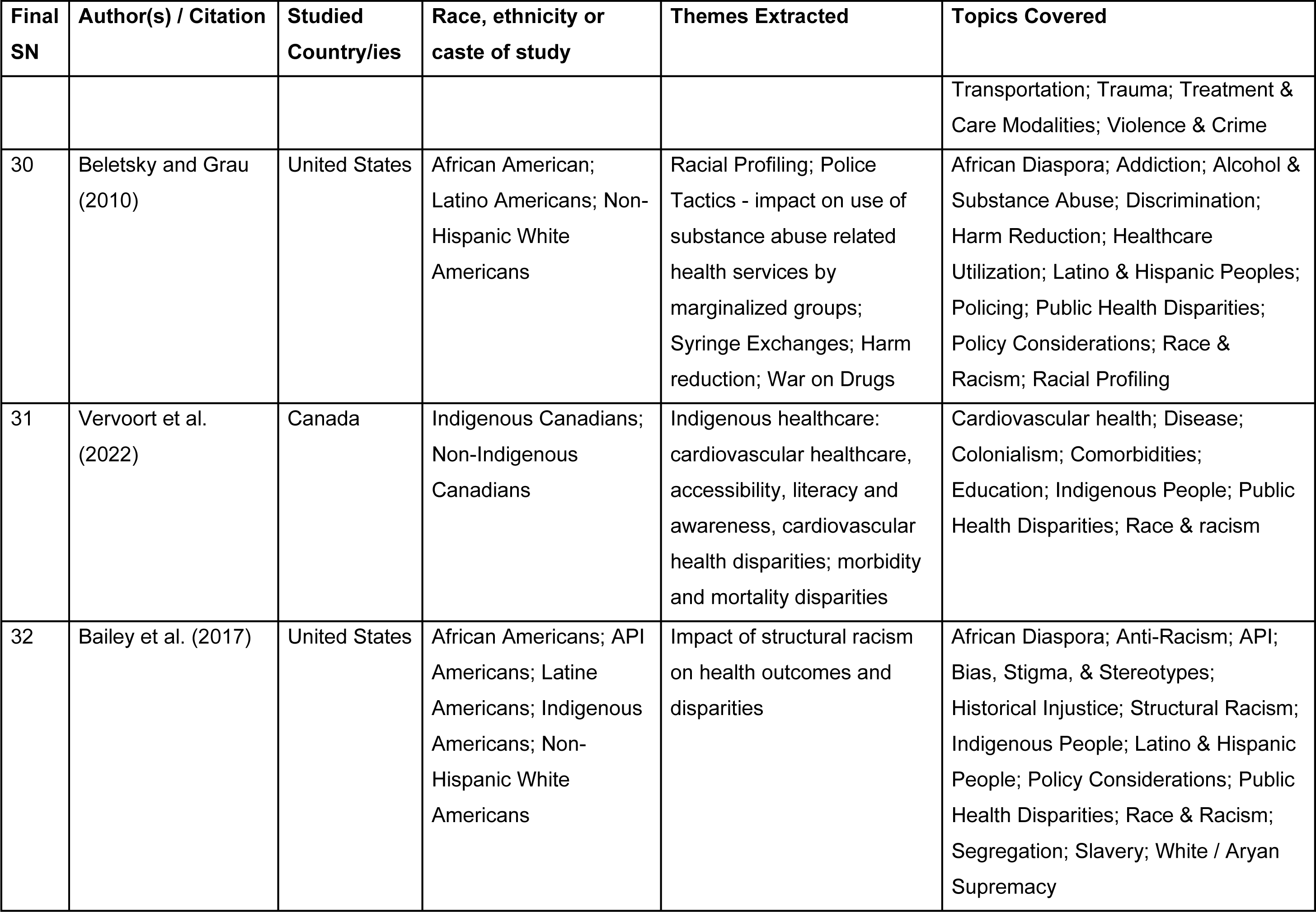
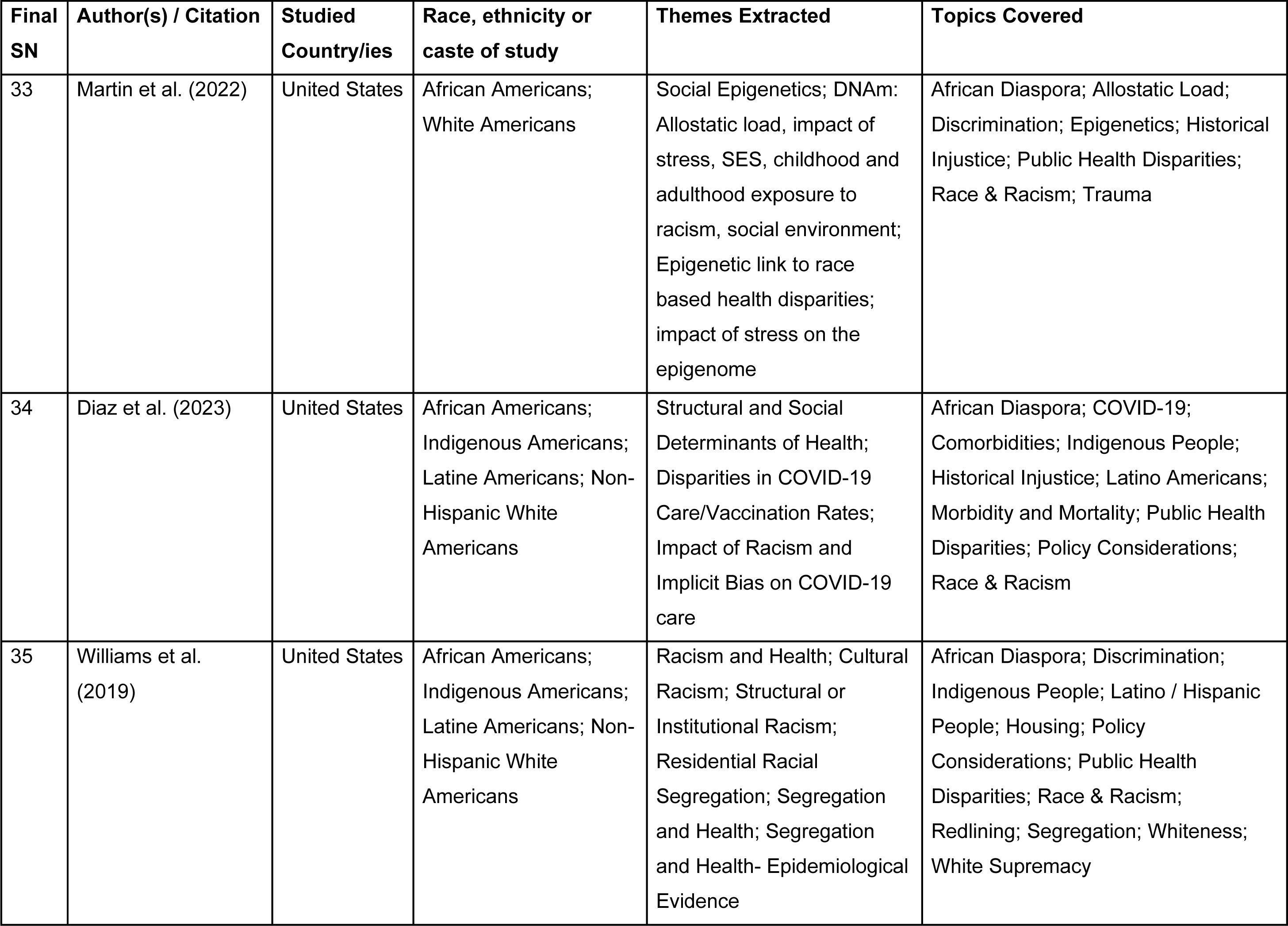
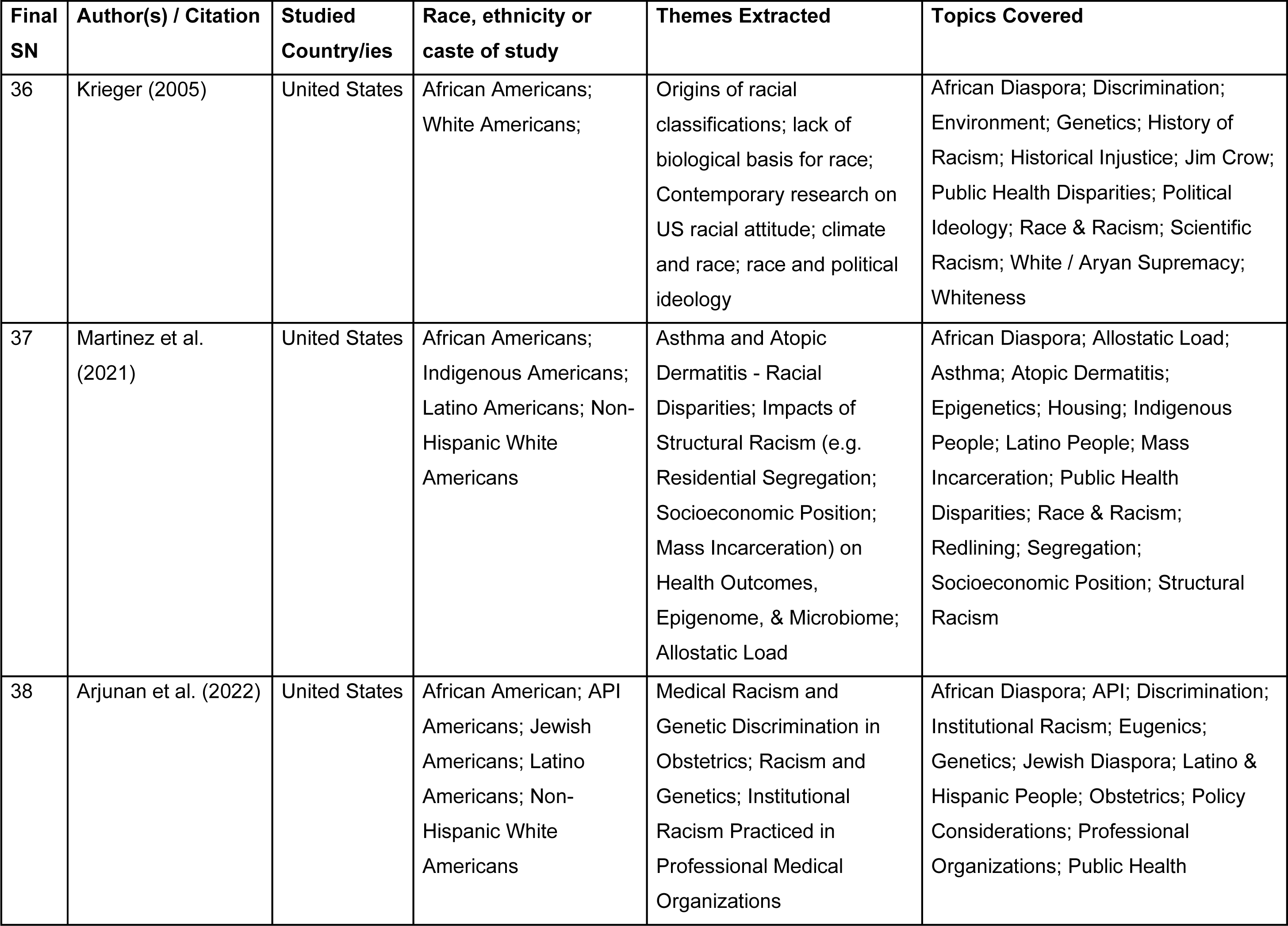
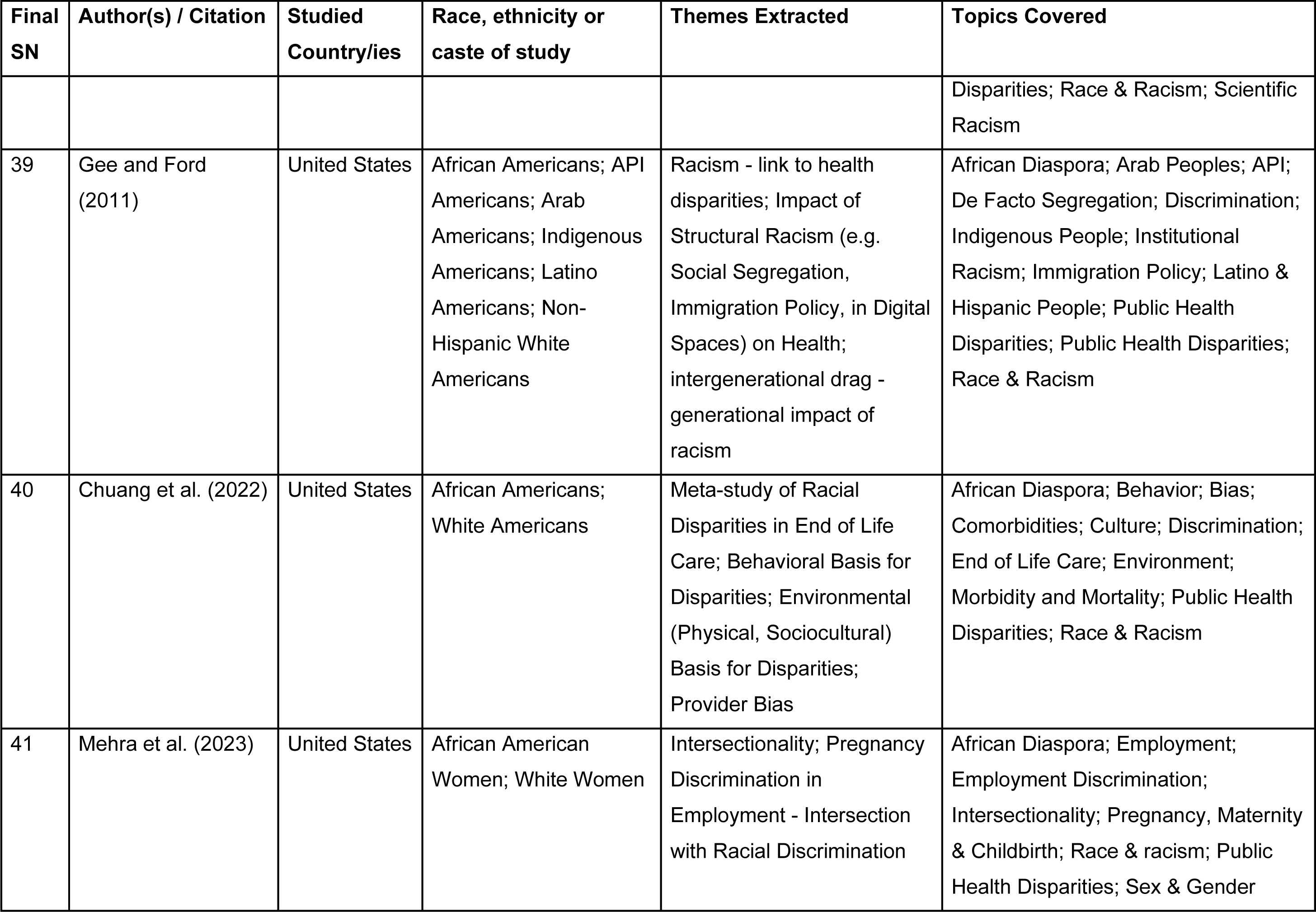
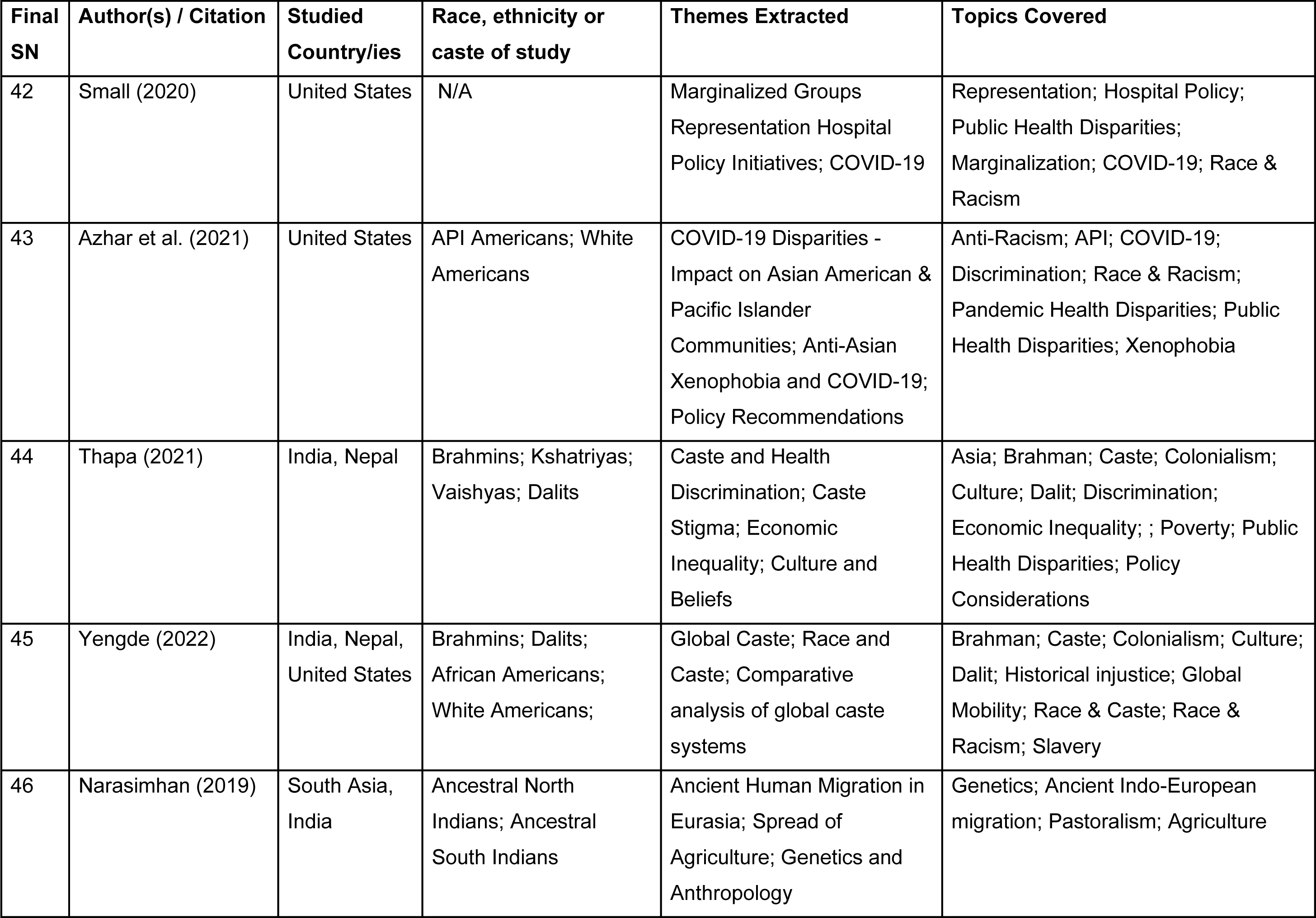
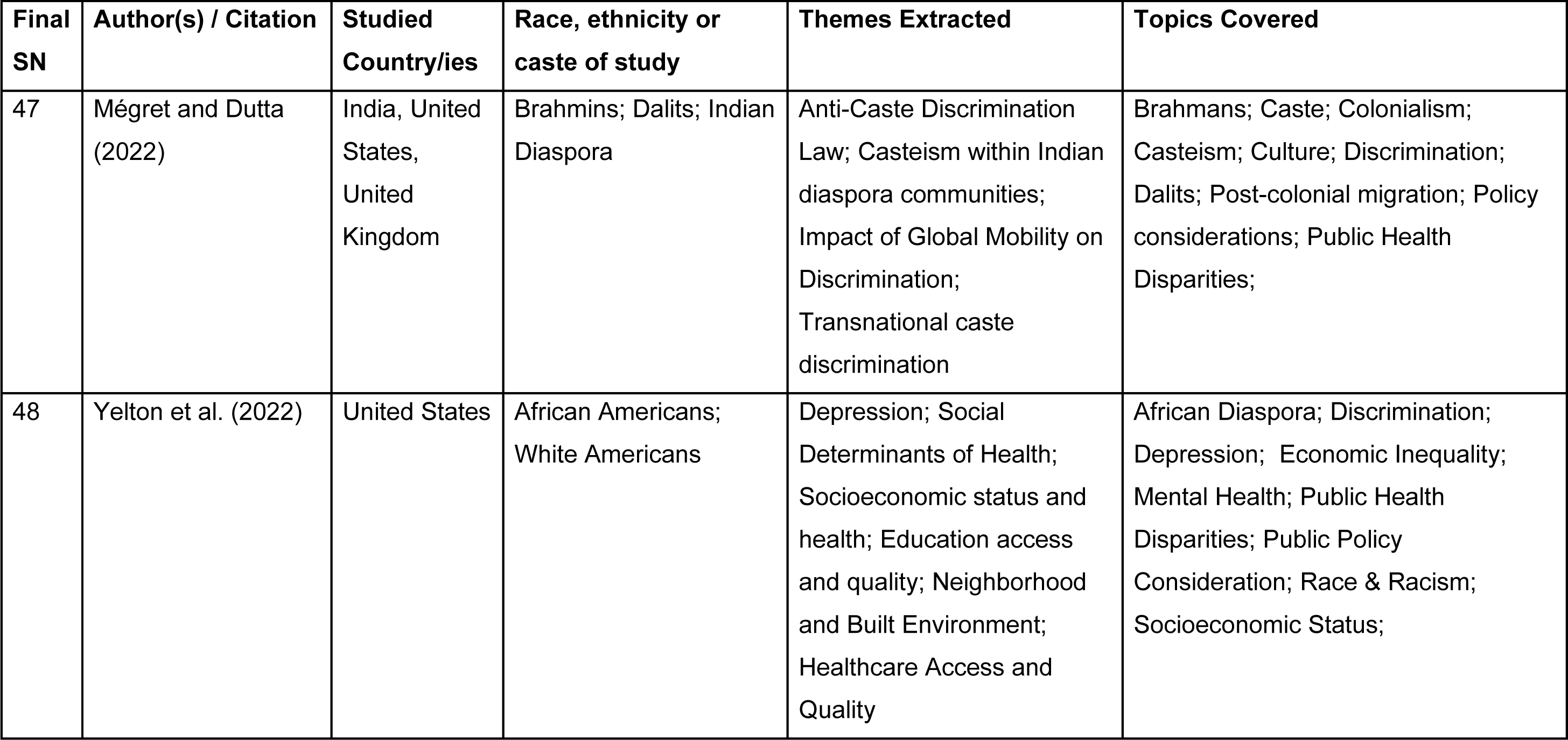
List of finally selected articles and their major characteristics.

| **Final SN** | **Author(s) / Citation** | **Studied Country/ies** | **Race, ethnicity or caste of study** | **Themes Extracted** | **Topics Covered** |
| --- | --- | --- | --- | --- | --- |
| 1 | Sweeting et al. (2023) | United States | African Americans;<br>White Americans | Parental preconception adversity; Epigenetic effects on health and disease; Structural racism as a root cause; Theoretical considerations | African Diaspora; Behavior; Epigenetics; Genetics; Pregnancy, Maternity, & Childbirth; Public Health Disparities; Race & Racism; Trauma |
| 2 | Hurd and Young (2023) | United States | African Americans;<br>Latino Americans;<br>Native Americans;<br>White Americans | Education, clinical child and adolescent psychology; Global Diversity Equity & Inclusion (DEI); decentering whiteness; discrimination (overrepresentation in punitive measures); Healthcare: access, clinicians' bias, representation among providers.; Mental health; Policy implications; Research needs; Socio-economic factors | African Diaspora; Anti-Racism; Behavior; Bias, Stigma, & Stereotypes; Care Access & Delays; DEI; Discrimination; Education; Indigenous People; Latino & Hispanic Peoples; Mental Health; Pediatric Care; People of Color (POC); Policy Considerations; Poverty; Race & Racism; Representation; Trauma; Treatment & Care Modalities; White / Aryan Supremacy; Whiteness |
| 3 | Subica and Link (2022) | United States | African Americans;<br>Native Americans;<br>White Americans | Cultural / historical trauma; Cultural trauma- mechanism of affecting health | African Diaspora; Culture; Discrimination; Historical Injustice; Housing; Indigenous People; Policy |

| Final SN | Author(s) / Citation | Studied Country/ies | Race, ethnicity or caste of study | Themes Extracted | Topics Covered |
| --- | --- | --- | --- | --- | --- |
|  |  |  |  | disparities; Fundamental cause theory | Considerations; Public Health Disparities; Race & Racism; Transportation; Trauma |
| 4 | Zhou and Wodtke (2019) | United States | Economic Class | Income class stratification and racial class stratification; Resource distribution - social stratification decline; Resource distribution - Socio-economic inequality | Class & Socioeconomic Status; Economic Inequality; Occupation; Race & Racism |
| 5 | Soled et al (2021) | United States | African Americans; Native Americans; White Americans | Reparations; Role of medicine and pseudo-science | African Diaspora; Care Access & Delays; Culture; Discrimination; Historical Injustice; Housing; Indigenous People; Policy Considerations; Public Health Disparities; Race & Racism; Trauma |
| 6 | Jacoby et al. (2018) | United States (Philadelphia) | African Americans; Latino Americans; White Americans | Crime; Gun Violence; Present day impacts of historical discrimination; Racial health disparities; Redlining | African Diaspora; Bias, Stigma, & Stereotypes; Discrimination; Ecology; Gun Violence; Historical Injustice; Housing; Economic Inequality; Latino & Hispanic Peoples; Policy Considerations; Race & Racism; Segregation; Violence & Crime |
| 7 | Tiedt and Brown (2014) | United States | African Americans;<br>Latino Americans;<br>White Americans | Historical trauma and present day discrimination as a source of chronic stress in Native Americans; Impact of stress (allostatic load) on diabetes rates | African Diaspora; Bias, Stigma, & Stereotypes; Care Access & Delays; Discrimination; Ecology; Gun Violence; Historical Injustice; Economic Inequality; Latino & Hispanic Peoples; Policy Considerations; Race & Racism; Segregation; Violence & Crime |
| 8 | Keyes (2009) | United States | African Americans;<br>White Americans | Health outcome impacts - coping and resilience; Health outcome impacts - mental health extent of discrimination; Health outcome impacts - morbidity and mortality; Health outcome impacts - risk factors | African Diaspora; Alcohol & Substance Abuse; Class & Socioeconomic Status; Comorbidities; Discrimination; Education; Economic Inequality; Mental Health; Occupation; Public Health Disparities; Race & Racism; Segregation; Trauma; Violence & Crime |
| 9 | Goodman et al (2017) | Canada (Vancouver) | Indigenous Canadians;<br>White Canadians | Indigenous health care: experience and outcomes; risk factors (alcohol), racially based adverse treatment; health outcomes, stereotypes and stigma; interrelated stigma and | Alcohol & Substance Abuse; Anti-Racism; Bias, Stigma, & Stereotypes; Care Access & Delays; Colonialism; Communication; Discrimination; Education; Economic Inequality; Indigenous People; Intersectionality; Mental Health; Policy Considerations; |

| Final SN | Author(s) / Citation | Studied Country/ies | Race, ethnicity or caste of study | Themes Extracted | Topics Covered |
| --- | --- | --- | --- | --- | --- |
|  |  |  |  | intersectional approaches; patients' stories | Poverty; Public Health Disparities; Race & Racism; Representation; Sexually Transmitted Infections; Trauma; Treatment & Care Modalities; Violence & Crime; White / Aryan Supremacy |
| 10 | McKinney et al. (2020) | United States | African Americans; Asian & Pacific Islanders (API); Latine Americans; Native Americans; Non-Hispanic White Americans | Genetic variations - genetic testing, stigma, genetic counseling, distrust among minorities, cultural competency training; Social cognitive theory | African Diaspora; API; Behavior; Bias, Stigma, & Stereotypes; Cancer; Care Access & Delays; Communication; Cultural Competence; Discrimination; Genetics; Genetic Testing; Housing; Personal Beliefs; POC; Policy Considerations; Public Health Disparities; Race & Racism; Religion; Treatment & Care Modalities |
| 11 | Browne et al. (2016) | Canada | Indigenous Canadians; White Canadians | Indigenous health care- 10 strategies to optimize effectiveness of health care; Structural violence - population and public health- major determinant of the distribution and outcomes of social and health inequities | Alcohol & Substance Abuse; Anti-Racism; Bias, Stigma, & Stereotypes; Care Access & Delays; Colonialism; Discrimination; Historical Injustice; Indigenous People; Policy Considerations; Pregnancy, Maternity, Childbirth; Public Health Disparities; Race & Racism; Trauma |
| 12 | Parra-Cardona et al. (2019) | United States | Mexican Americans; Non-Hispanic White Americans | Cultural adaptation; Mental health disparities - Mental health services in Latino immigrant communities | Behavior; Communication; Cultural Competence; Discrimination; Education; Latino & Hispanic Peoples; Mental Health; Parenting; Policy Considerations; Public Health Disparities; Race & Racism |
| 13 | Parkman et al. (2015) | United States | Connecticut, Michigan, Ohio, and Oregon Residents | Genetic privacy; anti-genetic discrimination policy | Communication; Discrimination; Genetics; Genetic Testing; Policy Considerations; Treatment & Care Modalities |
| 14 | Shaw and Armin (2011) | United States | People of Color Generally; White Americans | Cultural adaptation; physician training; corporatization of social justice interventions | Communication; Cultural Competence; Education; Ethnicity & Nationality; POC; Policy Considerations; Public Health Disparities; Race & Racism; Treatment & Care Modalities |
| 15 | Geronimus et al. (2010) | United States | African American Women; White American Women | Black-White health disparities; Intersection fo race and gender discrimination; accelerated biological aging in black women | African Diaspora; Age & Aging; Alcohol & Substance Abuse; Allostatic Load; Bias, Stigma, & Stereotypes; Discrimination; Environment; Genetics; Housing; Poverty; Public Health Disparities; Race & Racism; Sex/Gender; Trauma |
| 16 | Holtzclaw Williams (2011) | United States | N/A | Government funding - impact on medical research; anti-genetic discrimination police | Bias, Stigma, & Stereotypes; Discrimination; Care Access & Delays; Disease; Disability; Genetics; Pharmaceuticals; Policy Considerations; Public Health Disparities; Rare Disease; Treatment & Care Modalities |
| 17 | Lee (2009) | United States | African Americans; API; Latino Americans; Native Americans; Non-Hispanic White Americans | use of race and ethnicity in biomedical research; lack of a biological basis for race; race as a proxy for socioeconomic status, culture, and history | African Diaspora; API; Biology; Biomedical Research; Genetics; Genetic Testing; Indigenous People; Latino & Hispanic Peoples; Policy Considerations; Public Health Disparities; Race & Racism; Scientific Racism |
| 18 | Sankar et al. (2004) | United States | African Americans, Latino Americans, Native Americans, Non-Hispanic White Americans | Link between racial discrimination, poverty and health disparities; genetic research; public health policy; scientific racism | African Diaspora; Care Access & Delays; Discrimination; Education; Employment & Labor; Genetics; Genetic Testing; Housing; Indigenous People; Latino & Hispanic Peoples; POC; Policy Considerations; Poverty; Public Health Disparities; Race & Racism |
| 19 | Farmer and Ferraro (2005) | United States | African Americans; White Americans | Health outcomes in Black Americans; SES - interaction of SES in racial health disparities- interaction with Education largest | African Diaspora; Class & Socioeconomic Status; Care Access & Delays; Discrimination; Education; Employment & Labor; Economic Inequality; Occupation; Poverty; Public Health Disparities; Race & Racism; Segregation; Treatment & Care Modalities |
| 20 | Browne (2017) | Australia, Canada, New Zealand, United States | Indigenous People; White Americans, Australians, Canadians, and Kiwis | Indigenous health care in the Anglophone western hemisphere; racially based adverse treatment; health outcomes, stereotypes; historical injustice; Socioeconomic factors; | Alcohol & Substance Abuse; Discrimination; Indigenous People; Race & racism; Care Access & Delays; Policy Considerations; Public Health Disparities; Bias, Stigma, & Stereotypes |
| 21 | Thapa (2014) | Nepal | Brahman; Kshatriya (Chhetri); Dalit; Vaishya | Caste system in Nepal; Systemic discrimination; Allostatic Load; Anti-Dalit discrimination; Diabetes; Cultural Narratives | Allostatic Load; Asia; Brahman; Caste; Chhetri; Dalit; Discrimination; Diabetes; Poverty; Vaishya; Public Health Disparities |
| 22 | Sims (2010) | United States | African American Women; White Americans | Health outcomes in Black Americans; Discrimination - | African Diaspora; Age & Aging; Discrimination; Sex/Gender; Healthcare Utilization (Access & |

| Final SN | Author(s) / Citation | Studied Country/ies | Race, ethnicity or caste of study | Themes Extracted | Topics Covered |
| --- | --- | --- | --- | --- | --- |
|  |  |  |  | impact on trust in healthcare providers | Delays); Personal Beliefs; Public Health Disparities; Race & Racism |
| 23 | Jackson (2008) | United States | N/A | Alternatives to Race as a model of human diversity; historical origin of racism | African Diaspora; Bias, Stigma, & Stereotypes; White Supremacy; Whiteness; Scientific Racism; Race & Racism |
| 24 | Kaplan (2010) | United States | African Americans; White Americans | Race as a folk category; Black- White health disparities; Lack of a biological basis for racial categories; Impact of racism on public health outcomes | African Diaspora; Allostatic Load; Biology; Scientific Racism; Race & Racism; White Supremacy; Public Health Disparities; Discrimination; Historical Injustice; diabetes; cancer |
| 25 | Powell-Young and Spruill (2013) | United States | African Americans | Genetic Discrimination; Minority representation among healthcare professionals; African American views of genetic testing/Research | African Diaspora; Discrimination; Education; Genetics; Genetic Research & Testing; Nurses; Healthcare Utilization; Public Health Disparities; Race & Racism; Personal Beliefs; Representation |
| 26 | Bloche (2004) | United States | People of Color, White Americans | Systemic Racism; Ignoring race based disparities in health care to justify public health policies that perpetuate race based disparities; Ideological interference with public health policy | Race & Racism; Discrimination; Public Health Disparities; Care Quality; Policy; White Supremacy |
| 27 | Cabassa (2003) | United States | N/A | Practitioner bias in mental health care; multidisciplinary approach to finding the root of mental health disparity | Practitioner bias; Public Health Disparities; Mental Health; Race & Racism |
| 28 | Giddings (2005) | United States | People of Color; White Americans | Transcultural nursing; Cultural adaptation by healthcare professionals; social consciousness; Gender roles in Healthcare; Marginalization; traditional constructs in nursing | Race & racism; Cultural Competence; Nursing; Sex & Gender; Public Health Disparities |
| 29 | Kohrt et al. (2009) | Nepal | Brahman; Chhetri; Dalit; Janajati | Caste based discrimination in Nepal; Mental health - stresses | Age & Aging; Asia; Brahman; Caste; Chhetri; Childcare; Culture; Dalit; Discrimination; Education; Employment & Labor; Environment; Ethnicity & Nationality; Economic Inequality; Janajati; Mental Health; Occupation; Poverty; Public Health Disparities; Policy Considerations; Religion; Sex/Gender; Social Support; |
|  |  |  |  |  | Transportation; Trauma; Treatment & Care Modalities; Violence & Crime |
| 30 | Beletsky and Grau (2010) | United States | African American; Latino Americans; Non-Hispanic White Americans | Racial Profiling; Police Tactics - impact on use of substance abuse related health services by marginalized groups; Syringe Exchanges; Harm reduction; War on Drugs | African Diaspora; Addiction; Alcohol & Substance Abuse; Discrimination; Harm Reduction; Healthcare Utilization; Latino & Hispanic Peoples; Policing; Public Health Disparities; Policy Considerations; Race & Racism; Racial Profiling |
| 31 | Vervoort et al. (2022) | Canada | Indigenous Canadians; Non-Indigenous Canadians | Indigenous healthcare: cardiovascular healthcare, accessibility, literacy and awareness, cardiovascular health disparities; morbidity and mortality disparities | Cardiovascular health; Disease; Colonialism; Comorbidities; Education; Indigenous People; Public Health Disparities; Race & racism |
| 32 | Bailey et al. (2017) | United States | African Americans; API Americans; Latine Americans; Indigenous Americans; Non-Hispanic White Americans | Impact of structural racism on health outcomes and disparities | African Diaspora; Anti-Racism; API; Bias, Stigma, & Stereotypes; Historical Injustice; Structural Racism; Indigenous People; Latino & Hispanic People; Policy Considerations; Public Health Disparities; Race & Racism; Segregation; Slavery; White / Aryan Supremacy |
| 33 | Martin et al. (2022) | United States | African Americans;<br>White Americans | Social Epigenetics; DNAm: Allostatic load, impact of stress, SES, childhood and adulthood exposure to racism, social environment; Epigenetic link to race based health disparities; impact of stress on the epigenome | African Diaspora; Allostatic Load; Discrimination; Epigenetics; Historical Injustice; Public Health Disparities; Race & Racism; Trauma |
| 34 | Diaz et al. (2023) | United States | African Americans;<br>Indigenous Americans;<br>Latine Americans; Non-Hispanic White Americans | Structural and Social Determinants of Health; Disparities in COVID-19 Care/Vaccination Rates; Impact of Racism and Implicit Bias on COVID-19 care | African Diaspora; COVID-19; Comorbidities; Indigenous People; Historical Injustice; Latino Americans; Morbidity and Mortality; Public Health Disparities; Policy Considerations; Race & Racism |
| 35 | Williams et al. (2019) | United States | African Americans;<br>Indigenous Americans;<br>Latine Americans; Non-Hispanic White Americans | Racism and Health; Cultural Racism; Structural or Institutional Racism; Residential Racial Segregation; Segregation and Health; Segregation and Health- Epidemiological Evidence | African Diaspora; Discrimination; Indigenous People; Latino / Hispanic People; Housing; Policy Considerations; Public Health Disparities; Race & Racism; Redlining; Segregation; Whiteness; White Supremacy |
| 36 | Krieger (2005) | United States | African Americans;<br>White Americans; | Origins of racial classifications; lack of biological basis for race; Contemporary research on US racial attitude; climate and race; race and political ideology | African Diaspora; Discrimination; Environment; Genetics; History of Racism; Historical Injustice; Jim Crow; Public Health Disparities; Political Ideology; Race & Racism; Scientific Racism; White / Aryan Supremacy; Whiteness |
| 37 | Martinez et al. (2021) | United States | African Americans;<br>Indigenous Americans;<br>Latino Americans; Non-Hispanic White Americans | Asthma and Atopic Dermatitis - Racial Disparities; Impacts of Structural Racism (e.g. Residential Segregation; Socioeconomic Position; Mass Incarceration) on Health Outcomes, Epigenome, & Microbiome; Allostatic Load | African Diaspora; Allostatic Load; Asthma; Atopic Dermatitis; Epigenetics; Housing; Indigenous People; Latino People; Mass Incarceration; Public Health Disparities; Race & Racism; Redlining; Segregation; Socioeconomic Position; Structural Racism |
| 38 | Arjunan et al. (2022) | United States | African American; API Americans; Jewish Americans; Latino Americans; Non-Hispanic White Americans | Medical Racism and Genetic Discrimination in Obstetrics; Racism and Genetics; Institutional Racism Practiced in Professional Medical Organizations | African Diaspora; API; Discrimination; Institutional Racism; Eugenics; Genetics; Jewish Diaspora; Latino & Hispanic People; Obstetrics; Policy Considerations; Professional Organizations; Public Health |

| Final SN | Author(s) / Citation | Studied Country/ies | Race, ethnicity or caste of study | Themes Extracted | Topics Covered |
| --- | --- | --- | --- | --- | --- |
|  |  |  |  |  | Disparities; Race & Racism; Scientific Racism |
| 39 | Gee and Ford (2011) | United States | African Americans; API Americans; Arab Americans; Indigenous Americans; Latino Americans; Non-Hispanic White Americans | Racism - link to health disparities; Impact of Structural Racism (e.g. Social Segregation, Immigration Policy, in Digital Spaces) on Health; intergenerational drag - generational impact of racism | African Diaspora; Arab Peoples; API; De Facto Segregation; Discrimination; Indigenous People; Institutional Racism; Immigration Policy; Latino & Hispanic People; Public Health Disparities; Public Health Disparities; Race & Racism |
| 40 | Chuang et al. (2022) | United States | African Americans; White Americans | Meta-study of Racial Disparities in End of Life Care; Behavioral Basis for Disparities; Environmental (Physical, Sociocultural) Basis for Disparities; Provider Bias | African Diaspora; Behavior; Bias; Comorbidities; Culture; Discrimination; End of Life Care; Environment; Morbidity and Mortality; Public Health Disparities; Race & Racism |
| 41 | Mehra et al. (2023) | United States | African American Women; White Women | Intersectionality; Pregnancy Discrimination in Employment - Intersection with Racial Discrimination | African Diaspora; Employment; Employment Discrimination; Intersectionality; Pregnancy, Maternity & Childbirth; Race & racism; Public Health Disparities; Sex & Gender |
| 42 | Small (2020) | United States | N/A | Marginalized Groups<br>Representation Hospital<br>Policy Initiatives; COVID-19 | Representation; Hospital Policy;<br>Public Health Disparities;<br>Marginalization; COVID-19; Race & Racism |
| 43 | Azhar et al. (2021) | United States | API Americans; White Americans | COVID-19 Disparities -<br>Impact on Asian American & Pacific Islander Communities; Anti-Asian Xenophobia and COVID-19; Policy Recommendations | Anti-Racism; API; COVID-19; Discrimination; Race & Racism; Pandemic Health Disparities; Public Health Disparities; Xenophobia |
| 44 | Thapa (2021) | India, Nepal | Brahmins; Kshatriyas; Vaishyas; Dalits | Caste and Health<br>Discrimination; Caste Stigma; Economic Inequality; Culture and Beliefs | Asia; Brahman; Caste; Colonialism; Culture; Dalit; Discrimination; Economic Inequality; ; Poverty; Public Health Disparities; Policy Considerations |
| 45 | Yengde (2022) | India, Nepal, United States | Brahmins; Dalits; African Americans; White Americans; | Global Caste; Race and Caste; Comparative analysis of global caste systems | Brahman; Caste; Colonialism; Culture; Dalit; Historical injustice; Global Mobility; Race & Caste; Race & Racism; Slavery |
| 46 | Narasimhan (2019) | South Asia, India | Ancestral North Indians; Ancestral South Indians | Ancient Human Migration in Eurasia; Spread of Agriculture; Genetics and Anthropology | Genetics; Ancient Indo-European migration; Pastoralism; Agriculture |
| 47 | Mégret and Dutta (2022) | India, United States, United Kingdom | Brahmins; Dalits; Indian Diaspora | Anti-Caste Discrimination Law; Casteism within Indian diaspora communities; Impact of Global Mobility on Discrimination; Transnational caste discrimination | Brahmans; Caste; Colonialism; Casteism; Culture; Discrimination; Dalits; Post-colonial migration; Policy considerations; Public Health Disparities; |
| 48 | Yelton et al. (2022) | United States | African Americans; White Americans | Depression; Social Determinants of Health; Socioeconomic status and health; Education access and quality; Neighborhood and Built Environment; Healthcare Access and Quality | African Diaspora; Discrimination; Depression; Economic Inequality; Mental Health; Public Health Disparities; Public Policy Consideration; Race & Racism; Socioeconomic Status; |

### 3.1 Health disparities across various forms of racism and casteism

Race and caste-based discrimination pose a significant threat to population health within the context of the World Health Organization’s health definition. This issue manifests as structural racism or casteism, contributing to health disparities among marginalized groups. The various forms of racism or casteism can impact facets of health depending on the religious, cultural, social, and economic contexts in the countries or their regions. Our review focuses on health disparities in the United States, Canada, and South Asia, specifically Nepal. Below, we briefly outline characteristics of health disparities across the primary forms of racism and casteism prevalent in these regions.

#### 3.1.1 Health Disparities Impacting Black African Americans

Numerous studies have reported that racial discrimination against Black and African American people leads to health disparities in the United States (CDC, 2011; Gee and Ford, 2011; Bailey et al., 2017; Krieger, 2014; Sweeting, et al., 2023; Williams et al., 2019). In the 21^st^ century, new challenges emerged, including mass incarceration and racial disparities in education, jobs, housing, and healthcare. Disparities in healthcare and health outcomes have been well documented at the individual level but less known at the population level (Bailey et al., 2017). Some of the most striking patterns of health disparities reported as pertinent to structural racism can be specified. For instance, Black people have elevated rates of mental health conditions like anxiety and depression, and they experience a higher incidence of hypertension due to racism-induced chronic stress (Soled et al., 2021). However, due to racial marginalization, Black people are less likely to seek out or receive medical care than Whites (Farmer & Ferraro, 2004; Hard & Young, 2023). In 1937, the Home Owners Loan Corporation (HOLC) developed a map of Philadelphia, ensuring that Black people were more likely to reside in the disadvantaged neighborhoods. Decades later, Black Philadelphians faced five times the risk of being assaulted with a firearm compared to white residents (Jacoby et al., 2018). Black people, despite their higher resilience and coping ability, especially for mental disorders, are reported to have greater rates of physical disease morbidity than Whites (Keyes, 2009).

Socioeconomic status (SES) is generally deemed to play a prominent role in health disparities across populations (Rasali et al., 2016). However, it is crucial to note that controlling for SES and education does not fully eliminate these disparities. This is because the effects of racism extend beyond those standard measures of one’s life prospects. Evidence indicates that Black and White Americans living with the same SES and with the same level of education have very different life experiences (Kaplan, 2010). The ‘diminishing returns hypothesis,’ proposed by Farmer and Ferraro (2004) in the health context, suggests that the differences between Black and White people are most significant at the highest SES levels. This phenomenon is attributed to the interaction effect of race with SES, mainly education, showing that racial health disparities, such as in self-rated health, are most prominent at higher levels of SES.

#### 3.1.2 Health disparities impacting Indigenous Peoples

Native Americans and Pacific Islanders in the United States and Aboriginal populations encompassing the First Nations, Inuit, and Metis peoples in Canada are the Indigenous peoples who have a long history of experiencing colonial racial discrimination, including forced displacement, dispossession of land, cultural suppression or acculturations, cultural insensitivity, and stereotyping (Browne, 2017). Due to colonialism, Aboriginal peoples in Canada are particularly vulnerable to healthcare inequities and face significant disparities in health status, morbidity and mortality, and healthcare access compared to the non-Aboriginal population, with increasing evidence of health status inequities that require redressing as routine aspects of health care (Browne et al., 2016; Goodman et al., 2017).

#### 3.1.3 Racism-related health disparities impacting other ethnic groups

In the United States, Hispanic and Latino Americans, Asian Americans, Arab and Middle Eastern Americans, Jewish Americans, and other religion-specific minorities experience racial discrimination based on their languages, immigrant status, stereotyping, and other historical biases such as Chinese Exclusion Act, Japanese internment during World War II and anti-Semitic discrimination against Jews, racial profiling of other groups (Krieger, 2014; Subica & Link, 2021). In Canada, despite adopting the multiculturalism policy, the Visible Minorities comprising hundreds of ethnic nationalities face historical or immigration-related racial discrimination. Especially, Chinese and South Asians experience the memory of historical racial discrimination related to immigration (such as historical trauma due to the Chinese Immigration Act, 1885, known for the Chinese head tax and the Komagata Maru incident in 1914), while they continue to face structural discrimination, which was exacerbated during COVID-19 pandemic (Rasali et al., 2021). Chinese people were stigmatized for COVID-19 virus origin (Viladrich, 2021), and the South Asian population in British Columbia was increasingly associated with higher exposure to confirmed COVID-19 infection (Rasali et al., 2021) because of their living circumstances. All these forms of racial discrimination bring about intergenerational trauma and prejudices in employment or livelihood circumstances that result in chronic stressors leading to disparities in both healthcare access and health outcomes.

#### 3.1.4 Caste-based health disparities and the link between casteism, transnational casteism, and racism

In South Asia, particularly India and Nepal, the countries of caste origin, the traditional caste system with its four hierarchically tiered caste groups-Brahmin, Kshatriyas, Vaishya and Shudra, traditionally assigned by individuals’ birth, is the longest surviving system of social hierarchy and segregation that put the people, especially those who are now called Dalits, to the lowest stratum of the society, by historically relegating them to a position of inferiority (Mégret & Dutta, 2022). Recent anthropological and genomic research indicates that race and caste share common roots, stemming from the migration of Middle Eastern farmers and Great Steppe pastoralists to ancient India and Europe around 4,000 years ago (Reich, 2018; Narasimhan et al., 2019). In ancient India, the patterns of accumulation and concentrations of the agricultural production surplus by the dominant groups evolved into a hierarchical caste system (Reich, 2018; Narasimhan et al., 2019) through the initial segregation of people into groups by the knowledge and occupational skills gained. Eventually, it took a cultural and religious turn following the composition of Hindu scriptures *Rigveda* and *Manusmriti* (Mégret & Dutta, 2022) between 1200 and 1300 BCE.

A literature review of nine full articles conducted by Thapa et al. (2021) relates that individuals’ opportunities to access education, employment, and health care are negatively impacted by caste discrimination. Dalits experience the negative health impacts to a significantly greater extent due to both their caste status and poverty which put them at higher vulnerability to health risks, generally with denial of healthcare provisioning due to discrimination (Thapa et al., 2021).

##### Transnational casteism

The United States and Canada, continue to deal with the existing problem of race-based discrimination, due to the reinforcing and embodied structural racism (Krieger, 2014) as their domestic phenomenon. Mégret and Dutta (2022), studying from a legal perspective, shed light on a new phenomenon of transnational casteism in western countries, emerging as the migration of discriminatory practices through diasporic dispersion. The authors define the phenomenon as “the continuation of patterns of discrimination exported from the society of origin but manifesting themselves in a host country.” The authors relate that as casteism’s discriminatory practice was largely unknown to the societal systems in the host countries, the growing incidents of caste discrimination are facing a wave of legal challenges.

They highlighted the challenges the host states have faced, in terms of their laws and policies, in resolving the emerging problem due to difficulty in addressing through the existing discrimination legislation. The ability of dominant sectors of the diaspora to deny the existence of caste practices abroad reflects its power to set the narrative (Mégret and Dutta, 2022). The authors admitted that their frameworks of discrimination analysis remained greatly emphasizing a domestic context and its supranational supervision that leads to a situation of neglecting the extent to which transnational discrimination has increased through the migration patterns of intra-diasporic discrimination. From their case study based on the Indian diaspora, the authors make recommendations for further study on the origin, nature, and impact of transnational casteism, which can potentially change the concept of how discrimination plays out as well as how the law can address arbitration of cultural and political legacies from abroad to give voice to the discriminated group among diasporas. Considering the impacts of caste discrimination on health disparities reported from the countries of origin, transnational casteism is bound to have implications on health disparities in the host countries that deserve a discourse yet to be documented.

### 3.2 Theoretical underpinning

While structural racism as a determinant of health disparities in the United States and Canada is well documented as a complex issue (Bailey et al., 2017), caste discrimination from South Asia painted with cultural orientation despite its commonality with the former as global caste (Yengde, 2022) adds a further layer of complexity to our discourse. Considerations of theoretical underpinning are, therefore, essential to provide a systematic, informed, and comprehensive approach to understanding and addressing such a complex issue for advancing our knowledge that can deepen interdisciplinary insights with repeatability for better predictive power. In the literature, we have reviewed a few well-established theoretical and conceptual frameworks of racism, (as applied to caste discrimination as well), that have been reported in relation to health disparities. We consider four major theories that directly relate to structural racism or casteism that have causal relationships with the production of health disparities at the societal or population level transcending through generations.

Two seminal theoretical and conceptual analyses of society level health disparities in relation to race and ethnicity as their structural and systemic determinant have been well documented (Bailey et al., 2017; Mannor et al., 2021; White et al, 2020; Ford & Airhihenbuwa, 2010), explicitly, Krieger (2014)’s Ecosocial Theory, and Ford (2010)’s explanation of Critical Race Theory (CRT) from a public health perspective, which they refer to as Public Health Critical Race Praxis (PHCRP). Both guide the overarching contemporary worldview of structural racism and casteism, leading to both interpersonal and population-level health disparities. To provide a concrete, feasible, and tenable approach to improving population health equity, Bailey et al. (2017) suggested a focus on structural racism, referring to “the totality of ways of societies fostering racial discrimination through mutually reinforcing systems of various living factors (such as housing, education, employment), which, in turn, reinforces the discriminatory behaviors and impacts on the distribution of resources’’. Further, CRT, which was originally used in legal studies, is characterized by race consciousness, emphasizes contemporary societal dynamics and socially marginalized groups, and the praxis between research and practices (Ford & Airhihenbuwa, 2010). Ford and Airhihenbuwa (2010) applied the CRT to shift the paradigm for analysis and interventions to eliminate. It promotes health equity by responding to structural racism’s contemporary influence on health, health inequities, and research. Both theories could be extended to historical health disparities caused by the caste discrimination handed down for centuries. Proponents of both theories have already applied the frameworks to a wide range of racially disadvantaged ethnicities (e.g., Roma populations in eastern Europe). Another two theories, Historical Trauma Theory and Fundamental Cause Theory go further in explaining the mechanisms of how health disparities as damaging impacts are produced in racially disadvantaged groups. Historical trauma theory posits that disadvantaged groups with a unique history of race-based adverse life trauma experience physical, psychological, and economic disparities that persist through generations (Sotero, 2006; Tiedt & Brown, 2014; Sweeting et al., 2023). The psycho-spiritual impacts of multiple traumatic events such as forced relocation, forced assimilation and annihilation, termination, and discrimination transcending generations have resulted in post-traumatic stress disorder (PTSD) in Native Americans (Tiedt & Brown, 2014). Likewise, the experience of slavery, marginalization, discrimination, and systemic violence has caused African Americans to experience poorer health compared to whites throughout their lifespans and has had cascading effects across generations, contributing to widespread health disparities (Sweeting et al., 2023). On the other hand, Subica and Link (2022) outlined the fundamental cause theory, which posits that certain racial groups experience health disparities due to underlying socio-economic factors such as lower socio-economic status, stigma, and racism, restricting their ability to access health protective resources. The authors advocate applying the theory to any disadvantaged group, race, ethnicity (or caste), in describing their health disparities resulting from an impact of cultural trauma such as ongoing physical or psychological assault meted to them by an oppressive dominant group usurping another group’s cultural resources through force, threats of force, or oppressive policies (Subica & Link, 2022). However, the social factor in question must be able to impact multiple health outcomes in a population, embody access to flexible resources, and reinforce replaceable mechanisms over time, complying with structural racism or casteism. Additionally, Social Cognitive Theory posits a connection between one’s knowledge and beliefs and one’s actions and behaviors, indicating that evidence-based theoretical frameworks can be useful in assessing persistent genetic testing disparities among various populations. McKinney et al. (2020) used this theory to understand the psychological factors behind disparities in the use of genetic testing to detect cancer risks between Whites and other racialized populations. From an Indigenous population and public health perspective, structural discrimination has been understood as a major determinant of the distribution and outcomes of social and health inequities, creating the conditions that sustain the proliferation of health and social inequities (Browne et al., 2016). Defining structural discrimination as “structural violence,” Browne et al. (2016) referred to “the disadvantage and suffering that stems from the creation and perpetuation of structures, policies, and institutional practices that are innately unjust, because systemic exclusion and disadvantage are built into everyday social patterns and institutional processes.”

### 3.3 Issues of science

#### 3.3.1 Genetic variations

Genetic variations among human population groups are widely misunderstood due to the persistence of pseudo-scientific 17th-century theories that sought to divide populations into races or castes based on imagined biological differences. In 2005, Nancy Krieger clearly described the stormy “state of contemporary discourse on race, genetics, and health disparities in the United States over whether race is a biological or social construct, and also whether racial/ethnic disparities in health are due to either innate genetic differences or the biological impact of present and past histories of racial discrimination and economic deprivation, or both”. More recent scientific evidence has firmly established that the human race or caste is an anthropogenic socio-cultural construct driven by the dominant group of humans leading to socio-economic discrimination of marginalized groups and associated health disparities and do not hold a genetic basis to classify them as they are recognized in the society (Mégret & Dutta, 2022). Basing “race” on skin color alone is not tenable because skin color is not linked to a specific gene unique to individual races or castes. Instead, the phenotype of skin color is the expression of myriad combinations of multiple genes (Jackson, 2008). Over time, natural selection retains phenotypic traits best suited to the environmental conditions in a certain geographic area. The genetic basis of caste formation handed down from ancient India, on the other hand, was claimed merely on the notion of “purity” maintained through endogamy. However, recent anthropological and genomic research showed that all caste groups of India are descendants from admixtures of three ancient ancestries-Indigenous ancestries such as Dravidians, and two transmigrant ancestries from Anatolia and Steppe mountains, forming ancestral genomic clusters, namely Ancestral North Indians (ANI) and Ancestral South Indians (ASI), in a gradient of their varied proportions across the Indian populations (Narasimhan et al., 2019). Further evidence indicates that the socially identified categories of races comprise most genetic variations within, rather than between their populations (Kaplan, 2010, Reich, 2018), disproving the notion of a genetic basis for race or caste.

As the meaning, significance, and use of the race concept varied historically with economic, political, social, and cultural influences, some biomedical scientists may carry the belief that racial difference is biologically meaningful impacting on health differences across race and ethnicity groups (Lee, 2009). This stems from the science and medicine of early modern era that harbored the mythological notion of biological race, such as claiming the existence of “Black” genes inherited by Black individuals, resulting in their race as inherently inferior to White people, implying that behavior and physical features are solely determined by genetic characteristics, with impacts on differences in disease occurrences (Soled et al., 2021; Lee, 2009). For scientists and scholars who visualize race as a social construct, extant racial (or ethnic) categories can be used as a proxy for sociocultural, economic, and historical processes and experiences to capture behavioral and structural differences between racialized groups (Lee, 2009; Kaplan, 2010). Genetic variations do exist within and between given human populations, like in any organism. However, these variations do not conform to extant notions of racial categories and the associated racial health disparities (Lee, 2009). The presence or absence of numerous genes makes the differences in the causation of common or complex diseases, and they also may contribute to disparities in the disease prevalence across population groups; however, from the science of genetics, it is well understood that the causation of a disease is a result of a complicated process of interaction between genes and the environment (Sankar et al., 2004). Sankar et al. (2004) suggested that overemphasizing the contribution of genetics to the causation of health disparities could be misleading and even reinforce racial stereotyping, as considerable evidence exists indicating that health disparities resulted in the United States largely from “long-standing, pervasive racial and ethnic discrimination”, rather than the genetical effects. More recently, Martinez et al. (2021) also suggest that narrowly focusing on genetic variations undermines the complexity of interactions across biological and structural factors in the risk of causation of diseases such as atopic diseases, including asthma. Most human diseases, such as diabetes and hypertension, are polygenic and occur in association with a host of genetic risk factors, environmental risk factors, and social determinants of health, rather than tagged to a race or caste group (Thapa, 2014). A few relatively rare diseases like Sickle Cell disease associated with HbS allele commonly reported among Black people and Tay Sachs disease, thalassemia, commonly reported in Ashkenazi Jew community (Kaplan, 2010) are the results of gene mutations that occur under conditions of certain environmental exposures, originating from the malaria endemic regions, for a long time. Such diseases are inheritable diseases, as a single gene controls them. However, they are not inherently associated with the overall genomic composition of any race or ethnic group, rather the historical exposure by way of carriers of mutated genes, leading to the vulnerability of the people of socially constructed race or ethnicity.

#### 3.3.2 Genetic testing

The science of genetics brings light to the existential realities of humanity. However, with incomprehensive and biased usage of scientific knowledge in the past, genetics was taken for granted to support and justify the discrimination, criminalization, and institutionalization of individuals or groups of people categorized as certain races (McKinney et al., 2020). The historical injustices have created a mistrust in medical research, fear of discrimination and abuse, and reduced access to services like genetic counseling, leading to persistent disparities in the uptake of genetic services such as genetic testing in the United States (Parkman et al., 2014). Distrust among racialized communities continues to be a barrier to their use of genetic testing. However, evidence-based guidelines promoting genetic testing are increasingly available in some clinical settings (Parkman et al., 2014).

#### 3.3.3 Epigenetic changes

Martin et al. (2022), clarified that race is not biological; rather, it is a social construct used to organize people into a social hierarchy based on physical or imagined features. They also highlighted the importance of epigenetics, an emerging area of science that examines DNA Methylation (DNAm) of genes, including those responsible for stress and inflammation in response to adverse social environments in early life. Social epigenetics can shed light on biological pathways by which social experiences affect health outcomes, providing us with a plausible biological mechanism for health inequalities (Martin et al., 2022; Sweeting et al., 2023).

The intergenerational health consequences of historical trauma experienced among Holocaust survivors and Indigenous populations are well documented (Sweeting et al., 2023). The findings from the research evidence suggested that the historical intergenerational trauma caused by adversity, such as racial discrimination, can affect gene expressions, resulting in biological dysfunction through epigenetic alterations. Such adversity affects the immune, neuroendocrine, and cardiovascular systems, leading to epigenetic aging via the methylation of immune response and threat-related amygdala reactivity genes (Sweeting et al., 2023). Such epigenetic changes include exposure to racial discrimination associated with lower parasympathetic cardiac modulation as measured by heart-rate variability (Sweeting et al., 2023), low birth weight of babies resulting from an alteration in the DNAm associated with fetal malnutrition (Tiedt & Brown, 2014). Sweeting et al. (2023) highlighted that adversity experiences, such as the impact of racial discrimination during pregnancy, can contribute to behaviors in the offspring as the impact of the epigenetic changes in future generations such that these changes are associated with greater adiposity and body mass index in the offspring. We posit that at the societal level, such epigenetic impacts can cumulatively result in the behaviors or health outcomes disproportionately across populations, contributing to structural health disparities.

#### 3.3.4 Psychosocial stresses, allostatic load, and telomere shortening

Various chronic psychosocial stresses such as cultural trauma (Subica & Link, 2022) and social disadvantages (Sweeting et al., 2023) caused by pervasive racial or caste discrimination are well documented. Subica and Link (2022) emphasized the significant impact of cultural trauma from racial discrimination on affected groups, leading to social disadvantage, which can mediate pervasively psychosocial effects, ultimately resulting in mental and physical health disparities. Parra-Cardona et al. (2019), Soled et al. (2021), and by Keyes (2009) highlighted the consistent and independent association of chronic stress caused by racial discrimination with various health conditions, including anxiety, depression, hypertension, and physical disease. Farmer and Ferraro (2005), highlighting equity theory, suggested that feelings of unfairness meted from racial discrimination can lead to psychological distress, which in turn affects physical and mental health negatively on the affected people. Jocoby et al. (2018) shed light on the challenges faced by Black and Latino communities living in disadvantaged urban environments that are exposed to physical, social, and psychological consequences of violent crime and injuries, which further exacerbate negative impacts on their health and well-being. Likewise, the Native American population faces a multitude of chronic stressors, including political oppression, intergenerational trauma, and socioeconomic disparities that, in turn, influence behaviors in diet and physical activity, stress levels, and coping skills, contributing to health outcomes such as diabetes (Tiedt & Brown, 2014). Specific to caste discrimination in Nepal, an ethnographic study conducted by Kohrt et al. (2009) identified various mediators associated with caste-discrimination resulting in mental health disparities among caste groups. The authors posit that stressful life events, low resource ownership, low household income and lack of social support are such mediators that were significantly associated with health disparities.

Bailey et al. (2017) highlighted that growing body of scientific evidence is showing the association of experiences of racial discrimination at the interpersonal level with biomarkers of disease and wellbeing. These markers included higher allostatic load, inflammation, coronary artery calcification, dysregulation in cortisol, greater oxidative stress, and shorter telomere length (Bailey et al., 2017; Williams et al., 2019). The repeated activation of stress response systems triggered by the exposure to various chronic stresses based on racial or caste discrimination can produce “Allostatic load”, which is the “cumulative biological burden” to adapt to the demands for life’s survival and maintenance (Kaplan, 2010; Geronimus et al., 2010). In other words, exposure to chronic stresses followed by repeated activation of the stress response systems makes stress responses overused and weak resulting in an allostatic load as a compensatory measure which is the “weathering” or “wear and tear” in the body system. In this process, because of neuronal axis and hormonal dysfunctions, a loss of cortisol’s anti-inflammatory effects ensues increasing inflammation and oxidative stress, which in turn, pose an elevated risk of cardiovascular, immune, and metabolic dysfunction (Geronimus et al., 2010). Consequently, a high allostatic load negatively affects health. It is associated with increased hypertension, heart disease, increased risk of preterm birth and low birth weight, and poor self-reported health, potentially leading to the further intergenerational consequences of health impacts as reviewed by Kaplan (2010). For instance, Black Americans suffer from chronic stress throughout life, intergenerationally on an ongoing basis, ultimately resulting in earlier deaths (Kaplan, 2010) or higher mean allostatic load scores compared to Whites (Geronimus, 2010).

Telomeres, regions of repetitive DNA sequences that cap the end of a chromosome for stability, shorten as people age. It shortens with cell division due to oxidative stress breaking the DNA sequence. Geronimus et al. (2010) noted that oxidative stress, a biomarker measured using telomere length, is an important mechanism linking aging, psychosocial stress, biological stress activation, inflammation, and disease development. Geronimus et al. (2010) reported that middle-aged African American women have shorter telomeres than White women, and this difference was significantly associated with exposure to stressors. For instance, Black women aged 49-55 are biologically older than their White counterparts by an age of 7.5 years, with the former’s telomeres measuring 371 base pairs shorter than the latter’s. Stress and poverty account for 27% of this disparity. This was attributed to the fact that Black women are forced to adapt to repeated material, psychosocial, and environmental stressors throughout their lifetimes.

### 3.4 Social determinants of health and structural determinants of health disparities

Social Determinants of Health (SDOH), the non-medical social and economic factors or circumstances people face in life, overwhelmingly impact our lives and behavior and outweigh all other factors (known as risk factors). Therefore, reducing health inequalities depends on reducing socioeconomic and other inequalities (Marmot & Allen, 2014). The 2008 WHO Conceptual Framework for Action on Social Determinants of Health (WHO CSDH) looked at social determinants of health to explore the root cause of disparities and develop actionable opportunities for addressing health disparities (Marmot and Allen, 2014; Diaz et al., 2023), while the conventional aggregate classes may even be deemed by some scholars no longer important as a determinant of health in the modern society (Zhao and Wodtke, 2018). Many studies reviewed were consistent on several of these individual social determinants, such as race, employment, housing, income, education and healthcare (Sankar et al., 2004; Sims, 2010). More recent studies have explained that there are mutually reinforcing inequitable systems in these social determinants of health such as housing, education, employment, income, healthcare, criminal justice and so on, which are, in turn, reinforcing discriminatory beliefs, values, and distribution of resources that altogether lead to the risk of health disparities (Bailey et al., 2017), acting together (Williams, 2019, Subica and Link, 2021; Soled et al., 2021; Diaz et al., 2023) or interacting with each other, e.g. interactions between Black race and education, income and occupation (Farmer and Ferrano, 2005), resulting in health disparities at the population level. As the socioeconomic position determines education level, household income, assets, or employment (Martin et al., 2022), Soled et al. (2021) further put forward that inequitable morbidity and mortality, exacerbated during the COVID-19 pandemic across racial groups in the US showed how historical racism expressed in healthcare, housing, employment, and education has continued impacting on the society. It is also demonstrated that health benefits result from initiatives to improve household income, education, employment opportunities, housing, and neighborhood conditions (Williams et al., 2019).

There is consistent agreement among the studies in our review that racism and casteism need to be considered as structural determinants of health disparities among historically marginalized populations, specifically Black people and Native Americans in the United States, Indigenous peoples in Canada and Australia, and Dalits in Nepal (Browne et al., 2016; Bailey et al., 2017; Thapa et al., 2021; Martin et al., 2022; Diaz et al., 2023). Health disparities cannot be attributed simply to behavioral or cultural factors, “rather, they are embodied manifestations of the complex interplay of socio-historical, political, and economic determinants of health” (Browne, 2017; Goodman et al., 2017). Examining a host of disciplines and sectors, including medicine, public health, housing, and human resources in the US context, Bailey et al. (2017) posited that structural racism has a substantial role how the distributions of various social determinants of health are played out with respect to the population health. According to Bailey et al. (2017), structural racism refers to “the totality of ways in which societies foster racial discrimination through mutually reinforcing systems of housing, education, employment, earnings, benefits, credit, media, healthcare, and criminal justice”.

Understanding the structural nature of racism and its impacts on health, accounting for the impact of historical factors such as intergenerational trauma, we can visualize how multiple outcomes from its redressing would be derived in ripple effects. For example, Gee and Ford (2011) explained how the 1964 Civil Rights Act, which prohibited employment discrimination, changed the demographics of healthcare as more people of color entered medical school and nursing school and went on to serve previously underserved communities. Diaz et al. (2023) portrayed a framework of structural and social determinants of health contributing to the racial and ethnic-based disparities in healthcare access or outcomes in the United States, cutting across the healthcare continuum during the COVID-19 pandemic, when all the manifestations of unintended or untoward consequences were exacerbated universally, and also proposes potential areas for required action as well (Diaz et al., 2023).

### 3.5 Discussion on major issues in the review

Historically racism and casteism have occurred based on supposed race or caste differences, by descent, between dominant groups, such as Whites in the US and Canada and so-called higher castes in Nepal and marginalized groups, such as Black and Native Americans in the US, Indigenous peoples in Canada and Dalits in Nepal have a common root and consistent characteristics, re-confirming the Wilkerson’s integrated idea of global caste (Bassett, 2021; Yendge, 2022). The literature reviewed in our study supports the consensus that structural racism and casteism are major determinants of health disparities.

Krieger (2014)’s Ecosocial Theory and the Ford (2010)’s extension of CRT through a public health perspective (i.e., PHCRP) provide solid foundations for explaining the racism or casteism as the structural determinant of health disparities at the population level, while other researchers often view critical discussions on this topic as unwarranted (Browne, 2017). There exists no ambiguity in the fact that the color-blindness or caste-blindness of egalitarianism cannot fully address the persistent systemic bias or disadvantageousness which appears in a wide range of socio-cultural norms or practices-anything from subtle bias to grave harms (e.g. incidents of rape or killing of people), which could occur even when the perpetrators may not always realize their wrongdoings that are unacceptable deeds tantamount to a crime against humanity in the 21^st^ century.

The body of literature reviewed in our study is buttressed by the prior literature on the structural nature of racism and casteism and the implications for health inequities. The disparities in health outcomes are created systemically or structurally as complex consequences of long-standing historical racial or caste discrimination, mediated through commonly known SDoHs, passing through various stages from chronic stresses to genetic, epigenetic, physio-pathological changes to cause disproportionately the morbidities and mortalities among racialized populations. Based on both the fundamental cause theory and historical trauma theory (Sotero, 2006; Tiedt and Brown, 2014; Subica and Link, 2022; Sweeting et al., 2023), we can infer from the scientific evidence available at hand by the 21^st^ century that the table has turned upside down, as ‘the victim blaming game’ with the falsified claims of eugenics imposing the idea of racially marginalized people inheriting the inferior genes or biological characteristics is over; instead, the blame is now redirected to the dominant groups, or the state in most cases, as the perpetrators are responsible and accountable for causing those harms in a structural manner.

The body of 21st Century’s scientific knowledge has converged into a consensus with the findings that can be summarized from our review as: 1) genetic variations can naturally occur between and within population groups for various reasons other than the socially constructed races or castes (Kaplan, 2010; Martinez et al., 2021) ; 2) genetic testing can be useful means of improving the health of the populations provided their use is not prejudiced or stigmatized across socially constructed races or castes (Parkman et al., 2014; McKinney et al., 2020); 3) epigenetic changes occur due to intergenerational trauma and harms caused by structural racism (Tiedt and Brown, 2014; Sweeting et al., 2023); and 4) continuous exposure to chronic psychosocial stressors for a long period of time can cause ‘allostatic load’ on the discriminated people resulting in disproportionate rates of health outcomes, explaining the mechanism of pervasive structural health disparities across populations (Kohrt et al., 2009; Subica and Link, 2022; Sweeting et al., 2023).

Diaz et al. (2023)’s examination of the COVID-19 pandemic to visualize profound health disparities caused by structural and social determinants such as racism shed light on those determinants contributing to the health disparities in a holistic perspective and also provided the opportunity to recognize the areas of action. Clearly, the individual social determinants documented in the US study can serve as the proxies of racism, acting as fragmented parts of the whole structural determinant of the health disparities that are cross-cutting across the healthcare continuum or also perhaps at the upstream population level (Diaz et al., 2023). This indicated the potential that the manifestations of racism could have occurred with any combinations of all those social determinants at any stage of the healthcare continuum in a variety of complex scenarios, mediating the overall impacts of structural disparities, depending upon the various untoward circumstances and challenges faced by the marginalized people. The conceptual framework of upstream and proximal pathways of structural racism and its health impacts, adapted by Martinez et al. (2021) from the earlier work of other researchers, convincingly shows how racism and other discriminations, including casteism, acting as the structural determinants of health disparities, triggering various determinants of health to mediate through various pathways leading to biological embedding (e.g. psychosocial and physiological stresses, adaptive immune response, etc.) resulting in racial disparity outcomes such as in allergy and immune functioning). The authors further explained these pathways to demonstrate the mechanism of the biologic embedding.

Vervoort et al. (2022) narrated the vulnerability of Indigenous peoples in Canada being at higher risk of cardiovascular disease compared to non-Indigenous people, with multiple factors such as historical oppression, racism, healthcare biases, etc. These findings were consistent with structural violence suggested by Browne et al. (2016), who posit that racism and discrimination must be considered determinants of health for Indigenous peoples. Likewise, Thapa et al. (2021) explicitly posit that caste is a fundamental determinant of social exclusion and development, impacting wider determinants of health such as education, employment, income, and housing.

Overall, addressing structural determinants of health disparities may require targeting cultural trauma through policy changes or direct interventions (Subica & Link, 2022) for a way forward in rectifying the historical injustices through reparative policy or intervention programs that are race-conscious rather than race-blind (Soled et al., 2021).

This study was implemented with a short timeline for the completion of extraction and synthesis of cross-disciplinary knowledge as an initiation for providing a global perspective on racial and caste discrimination as a structural determinant of health disparities. We might have also missed a vast wealth of literature from India in our effort to include less studied jurisdiction of caste discrimination in Nepal as mandated by the research program, especially in relation with health disparities. For that reason, the search terms were squeezed to extract the essence of knowledge from our targeted interests rather than making it an exhaustive treatise from the vast areas of multiple disciplines from health sciences to social sciences. We see this as a limitation of our study that can be expanded in future studies along the lines of this research.

### 3.6 Reparative policy considerations

Several of our selected articles have discussed implications of research that would inform policy formation or suggested interventions through policy changes towards eliminating health disparities rooted to racism or casteism. A set of recommended reparative policy changes for research and interventions extracted from the articles reviewed are presented in Table 2.

**Table 2.**
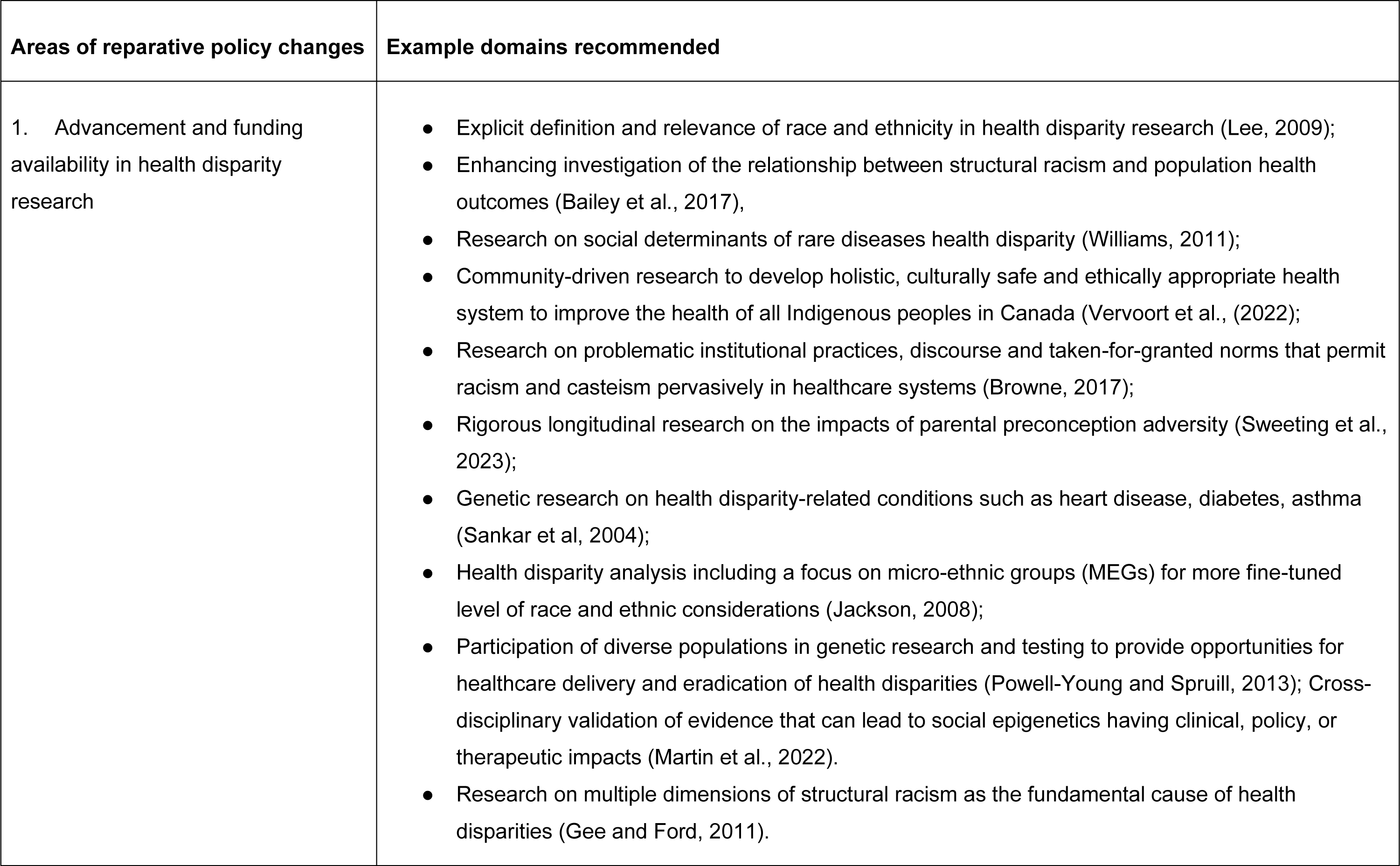
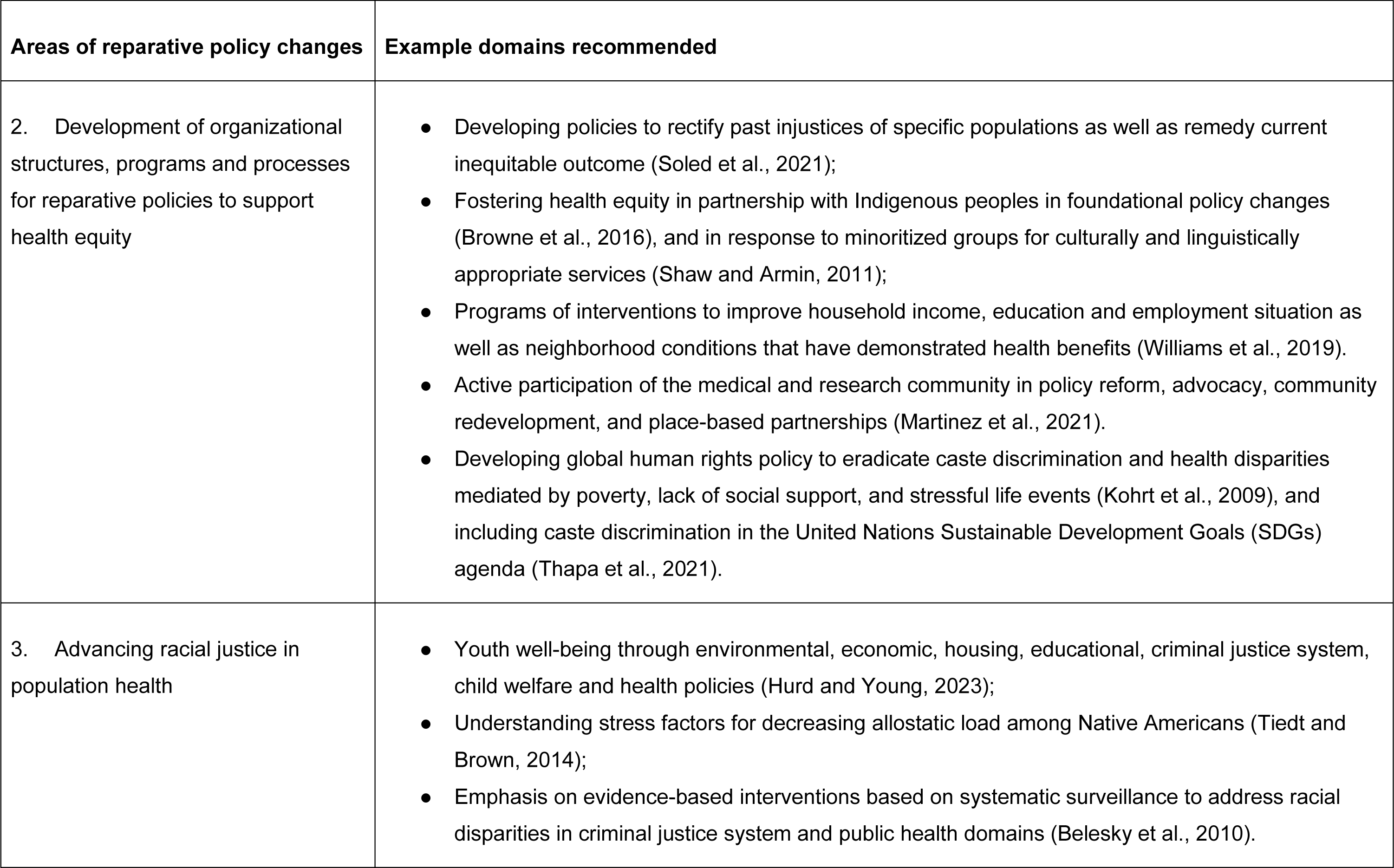
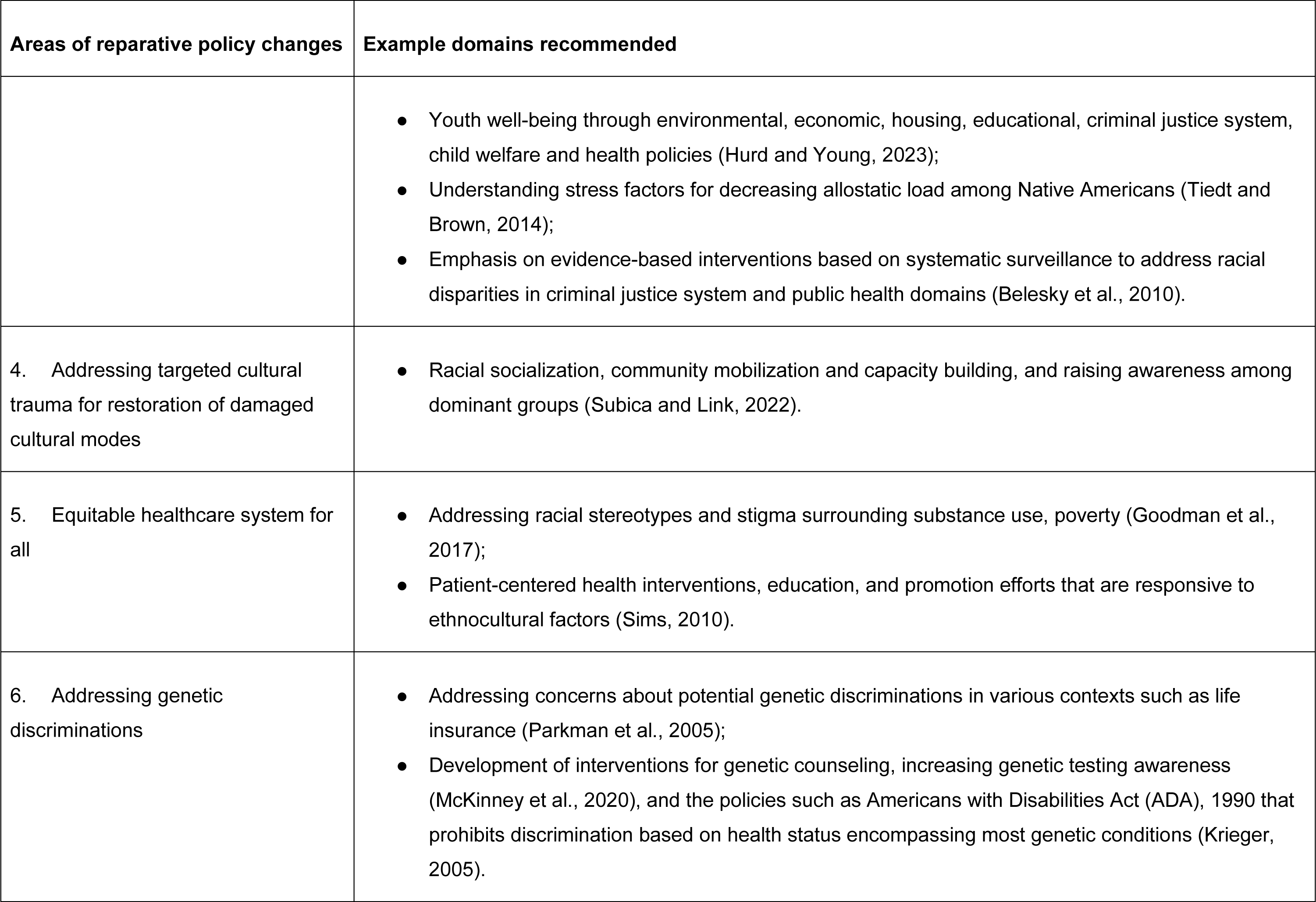
Major areas of reparative policy changes recommended for elimination of structural health disparities associated with racism and/or casteism, based on the articles reviewed.

| Areas of reparative policy changes | Example domains recommended |
| --- | --- |
| 1. Advancement and funding availability in health disparity research | • Explicit definition and relevance of race and ethnicity in health disparity research (Lee, 2009);<br>• Enhancing investigation of the relationship between structural racism and population health outcomes (Bailey et al., 2017),<br>• Research on social determinants of rare diseases health disparity (Williams, 2011);<br>• Community-driven research to develop holistic, culturally safe and ethically appropriate health system to improve the health of all Indigenous peoples in Canada (Vervoort et al., (2022);<br>• Research on problematic institutional practices, discourse and taken-for-granted norms that permit racism and casteism pervasively in healthcare systems (Browne, 2017);<br>• Rigorous longitudinal research on the impacts of parental preconception adversity (Sweeting et al., 2023);<br>• Genetic research on health disparity-related conditions such as heart disease, diabetes, asthma (Sankar et al, 2004);<br>• Health disparity analysis including a focus on micro-ethnic groups (MEGs) for more fine-tuned level of race and ethnic considerations (Jackson, 2008);<br>• Participation of diverse populations in genetic research and testing to provide opportunities for healthcare delivery and eradication of health disparities (Powell-Young and Spruill, 2013); Cross-disciplinary validation of evidence that can lead to social epigenetics having clinical, policy, or therapeutic impacts (Martin et al., 2022).<br>• Research on multiple dimensions of structural racism as the fundamental cause of health disparities (Gee and Ford, 2011). |
| 2. Development of organizational structures, programs and processes for reparative policies to support health equity | • Developing policies to rectify past injustices of specific populations as well as remedy current inequitable outcome (Soled et al., 2021);<br>• Fostering health equity in partnership with Indigenous peoples in foundational policy changes (Browne et al., 2016), and in response to minoritized groups for culturally and linguistically appropriate services (Shaw and Armin, 2011);<br>• Programs of interventions to improve household income, education and employment situation as well as neighborhood conditions that have demonstrated health benefits (Williams et al., 2019).<br>• Active participation of the medical and research community in policy reform, advocacy, community redevelopment, and place-based partnerships (Martinez et al., 2021).<br>• Developing global human rights policy to eradicate caste discrimination and health disparities mediated by poverty, lack of social support, and stressful life events (Kohrt et al., 2009), and including caste discrimination in the United Nations Sustainable Development Goals (SDGs) agenda (Thapa et al., 2021). |
| 3. Advancing racial justice in population health | • Youth well-being through environmental, economic, housing, educational, criminal justice system, child welfare and health policies (Hurd and Young, 2023);<br>• Understanding stress factors for decreasing allostatic load among Native Americans (Tiedt and Brown, 2014);<br>• Emphasis on evidence-based interventions based on systematic surveillance to address racial disparities in criminal justice system and public health domains (Belesky et al., 2010). |
|  | • Youth well-being through environmental, economic, housing, educational, criminal justice system, child welfare and health policies (Hurd and Young, 2023);<br>• Understanding stress factors for decreasing allostatic load among Native Americans (Tiedt and Brown, 2014);<br>• Emphasis on evidence-based interventions based on systematic surveillance to address racial disparities in criminal justice system and public health domains (Belesky et al., 2010). |
| 4. Addressing targeted cultural trauma for restoration of damaged cultural modes | • Racial socialization, community mobilization and capacity building, and raising awareness among dominant groups (Subica and Link, 2022). |
| 5. Equitable healthcare system for all | • Addressing racial stereotypes and stigma surrounding substance use, poverty (Goodman et al., 2017);<br>• Patient-centered health interventions, education, and promotion efforts that are responsive to ethnocultural factors (Sims, 2010). |
| 6. Addressing genetic discriminations | • Addressing concerns about potential genetic discriminations in various contexts such as life insurance (Parkman et al., 2005);<br>• Development of interventions for genetic counseling, increasing genetic testing awareness (McKinney et al., 2020), and the policies such as Americans with Disabilities Act (ADA), 1990 that prohibits discrimination based on health status encompassing most genetic conditions (Krieger, 2005). |

Many of the articles reviewed have focused on research gaps in supporting evidence-based policy changes leading to interventions (Lee, 2009: Williams, 2011; Gee and Ford (2011); Powell-Young and Spruill, 2013; Browne, 2017; Vervoort et al., 2022; Sweeting et al., 2023), while Bailey et al. (2017) even suggested that the health impacts of policy changes and interventions with potential for dismantling structural racism have been less studied. US NIH’s 1994 “inclusion mandate” brought women and minorities into clinical research; however, the operationalization of “race” and/or “ethnicity” was not explicit in 82% of the research articles reviewed by Lee (2009). Williams (2011) suggested that the US Health Disparity Research and Development Act was relevant to future research for rare disease policy development, while Powell-Young and Spruill (2013) posited that diverse populations’ participation in genetic research would lead to improving healthcare for the elimination of health disparities. Gee and Ford (2011) recommended more studies on disparities, especially considering the multiple dimensions of structural racism as fundamental causes of health disparities. Browne (2017) even showed the urgency of research to develop effective interventions for disrupting the processes problematic to providing the care for stigmatized people. Vervoort et al. (2022) suggested that community-driven research would help us better understand the gaps in holistic cardiovascular care so that healthcare and systems interventions could be made culturally safe and ethically appropriate for Indigenous peoples in Canada. Sweeting et al. (2023) pointed out the need for funding for “rigorous longitudinal research” necessary, especially in African Americans, for examining the negative health impacts of “intergenerational transmission of trauma “such as the impact of parental preconception adversity on offspring that could lead to the public health interventions.

Mental health is profoundly reported in the literature as one of the main areas of policy implications owing to the structural nature of extant health disparities linked to racism or casteism, especially among African American youth (Hurd and Young, 2023), Indigenous peoples of Canada and Native Americans (Tiedt & Brown, 2014; Goodman et al., 2017), and Dalits in Nepal (Kohrt et al., 2009). A few selected articles described policy implications for understanding stress factors in the Native American population (Tiedt & Brown, 2014), targeting intervention to reduce caste-based disparities in mental health such as in rural Nepal, addressing poverty, lack of social support, and stressful life events (Kohrt et al, 2009), and advancing mental health with racial justice (Hurd &Young, 2023).

Genetic discrimination has been historically a major issue of racism that led to the false claim of biological differences between races, as described in the preceding section. In the 21^st^ century, the policy reform is needed to address concerns among patients, research participants, and other stakeholders, including healthcare providers, especially about potential genetic discrimination in US life insurance (Parkman et al., 2015) and for the development of guides for genetic counseling to serve as an intervention to address causal genetic beliefs (McKinney et al., 2020). Longitudinal studies are suggested for formulating effective interventions to reduce health disparities through building evidence of the primary biological mechanism of DNAm providing social epigenetics knowledge that has clinical, policy and therapeutic impacts (Martin et al., 2022). Immigration policy is another area of structural racism, since the US’s exclusionary Immigration policy uses racial groups to reinforce social hierarchy, resulting in disparate health impacts (Gee and Ford, 2011). Williams et al. (2019) posit that anti-immigrant policies adversely affect population health, creating hostility towards immigrants and leading to their vulnerability. Other areas of policy implications that were drawn from the selected articles reviewed include the policy considerations in “geographically identified micro-ethnic groups for more nuanced and sensitive level analysis than race” (Jackson, 2008), in addressing health disparities through a patient-centered approach encompassing the outreach and capacity building efforts (Sims, 2010), and in emphasizing on the evidence-based interventions based on systemic surveillance of key criminal justice-related events that have health impacts (Beletsky et al., 2010).

## 4. Conclusion

In the cross-disciplinary review of literature from the 21st century, it becomes evident that race and caste stem from a shared socio-cultural construct dating back to the ancient times. Both have common characteristics of dominant groups oppressing the vulnerable groups in the society for ensuring dominant groups’ own socio-cultural and economic advantage over power, privilege, and resources. Racism and casteism evolved, and flourished, especially during the colonization period of the 17th century onwards reaching egregious levels of slavery, and dehumanizing discrimination such as segregation and untouchability. While the 21st century has witnessed strides in socio-political development fostering rising awareness aligning with human rights worldview and progressively realizing social change, there are many challenges in overcoming the structural nature of racism and casteism that serve as the structural determinant of health disparities across populations. Accordingly, the pseudo-scientific genetic discrimination, epigenetic effects of intergenerational trauma, disproportionate rates of various disease conditions caused by ‘allostatic load’ (e.g., diabetes, hypertension, mental health setbacks), and lingering social stigma, are still extant, with several risk factors and social determinants of health acting as mediators. The structural or societal level health disparities were consistently reported from all three case study countries, the US, Canada, and Nepal. By itself, the current approach to mitigating risk factors by focusing on the social determinants of health is inadequate for eliminating the impacts of structural racism and casteism. Reparative policy changes and appropriate evidence-based interventions informed by cross-disciplinary scientific research are imperative to achieve desired impacts towards health equity at the societal level.

## Authors’ contributions

Conceptualization: DPR, KTS; Data curation: DPR, BMW; Formal Analysis: DPR, BMW, Funding acquisition: DPR, KTS; Investigation: DPR, BMW, FAA; Methodology: DPR; Project Administration: DPR, KTS; Resources: DPR, KTS, DK; Software: DPR, BMW; Supervision: DPR, KTS, DK; Validation: DPR, FAA; KTS, WDO, SJ, CLF; Visualization: DPR, BMW; Writing-original draft-DPR; Writing – review & editing: DPR, BW, FAA, DK, KTS, WDO, SJ, CLF.

## Data Availability

All data produced in the present study are available upon reasonable request to the authors

## References

Arjunan, A., Darnes, D.R., Sagaser, K.G., Svenson, A.B., 2022. Addressing Reproductive Healthcare Disparities through Equitable Carrier Screening: Medical Racism and Genetic Discrimination in United States’ History Highlights the Needs for Change in Obstetrical Genetics Care. Societies 12, 33. 10.3390/soc12020033

Azhar, S., Farina, A., Alvarez, A.R.G., Klumpner, S., 2022. Asian in the Time of COVID-19: Creating a Social Work Agenda for Asian American and Pacific Islander Communities. Social Work 67, 58–68. 10.1093/sw/swab044

Bailey, Z.D., Krieger, N., Agénor, M., Graves, J., Linos, N., Bassett, M.T., 2017. Structural Racism and Health Inequities in the USA: Evidence and Interventions. The Lancet 389, 1453–1463. 10.1016/s0140-6736(17)30569-x

Bassett, M.T., 2021. Racism and Caste: Lessons for the USA. The Lancet 397, 187–188. 10.1016/s0140-6736(20)32762-8

Beletsky, L., Grau, L.E., White, E., Bowman, S., Heimer, R., 2010. The roles of law, client race and program visibility in shaping police interference with the operation of US syringe exchange programs. Addiction 106, 357–365. 10.1111/j.1360-0443.2010.03149.x

Bloche, M.G., 2004. Health Care Disparities — Science, Politics, and Race. New England Journal of Medicine 350, 1568–1570. 10.1056/nejmsb045005

Blocker, K., Hallford, H.G., McElfish, P., Danylchuk, N.R., Dean, L.W., 2020. Eliciting culturally and medically informative family health histories from Marshallese patients living in the United States. Journal of Genetic Counseling 29, 440–450. 10.1002/jgc4.1249

Browne, A.J., 2017. Moving beyond description: Closing the health equity gap by redressing racism impacting Indigenous populations. Social Science & Medicine 184, 23–26. 10.1016/j.socscimed.2017.04.045

Browne, A.J., Varcoe, C., Lavoie, J., Smye, V., Wong, S.T., Krause, M., Tu, D., Godwin, O., Khan, K., Fridkin, A., 2016. Enhancing health care equity with Indigenous populations: evidence-based strategies from an ethnographic study. BMC Health Services Research16. 10.1186/s12913-016-1707-9

Cabassa, L.J., 2003. Integrating cross-cultural psychiatry into the study of mental health disparities. American Journal of Public Health 93, 1034–1034. 10.2105/ajph.93.7.1034

CDC, 2011. CDC Health Disparities and Inequalities Report — United States, 2011. Centers for Disease Control and Prevention, US Department of Health and Human Services, Atlanta, GA.

Chuang, E., Yu, S., Georgia, A., Nymeyer, J., Williams, J., 2022. A Decade of Studying Drivers of Disparities in End-of-Life care for Black Americans: Using the NIMHD Framework for Health Disparities Research to Map the Path Ahead. Journal of Pain and Symptom Management 64, e43–e52. 10.1016/j.jpainsymman.2022.03.017

de Gobineau, A., 1915. The Inequality of Human Races, 1st English Edition. ed. William Heinemann, London.

Diaz, A.A., Thakur, N., Celedón, J.C., 2023. Lessons Learned from Health Disparities in Coronavirus Disease-2019 in the United States. Clinics in Chest Medicine 44, 425–434. 10.1016/j.ccm.2022.11.021

Dor Bahadur Bista, 1991. Fatalism and Development. Orient Longman.

Farmer, M.M., Ferraro, K.F., 2005. Are racial disparities in health conditional on socioeconomic status? Social Science & Medicine 60, 191–204. 10.1016/j.socscimed.2004.04.026

Ford, C.L., Airhihenbuwa, C.O., 2010. Critical Race Theory, Race Equity, and Public Health: Toward Antiracism Praxis. American Journal of Public Health 100, S30–S35. 10.2105/ajph.2009.171058

GBD US Health Disparities Collaborators, 2022. Life expectancy by county, race, and ethnicity in the USA, 2000–19: a systematic analysis of health disparities. The Lancet 400, 25–38. 10.1016/S0140-6736(22)00876-5

Gee, G.C., Ford, C.L., 2011. Structural racism and health inequities. Du Bois Review: Social Science Research on Race 8, 115–132. 10.1017/s1742058×11000130

Geronimus, A.T., Hicken, M.T., Pearson, J.A., Seashols, S.J., Brown, K.L., Cruz, T.D., 2010. Do US Black Women Experience Stress-Related Accelerated Biological Aging? Human nature (Hawthorne, N.Y.) 21, 19–38. 10.1007/s12110-010-9078-0

Giddings, L.S., 2005. A Theoretical Model of Social Consciousness. Advances in Nursing Science 28, 224–239. 10.1097/00012272-200507000-00005

Goodman, A., Fleming, K., Markwick, N., Morrison, T., Lagimodiere, L., Kerr, T., 2017. “They treated me like crap and I know it was because I was Native”: The healthcare experiences of Aboriginal peoples living in Vancouver’s inner city. Social Science & Medicine 178, 87–94. 10.1016/j.socscimed.2017.01.053

Holtzclaw Williams, P., 2011. Policy Framework for Rare Disease Health Disparities. Policy, Politics, & Nursing Practice 12, 114–118. 10.1177/1527154411404243

Hurd, N.M., Young, A., 2023. Introduction to the Special Issue: Advancing Racial Justice in Clinical Child and Adolescent Psychology. Journal of Clinical Child and Adolescent Psychology 52, 311–327. 10.1080/15374416.2023.2202255

Jackson, F.L.C., 2008. Ethnogenetic layering (EL): an alternative to the traditional race model in human variation and health disparity studies. Annals of Human Biology 35, 121–144. 10.1080/03014460801941752

Jacoby, S.F., Dong, B., Beard, J.H., Wiebe, D.J., Morrison, C.N., 2018. The enduring impact of historical and structural racism on urban violence in Philadelphia. Social Science & Medicine 199, 87–95. 10.1016/j.socscimed.2017.05.038

Kaplan, J.M., 2010. When Socially Determined Categories Make Biological Realities. Monist 93, 281–297. 10.5840/monist201093216

Keyes, C.L.M., 2009. The Black-White Paradox in Health: Flourishing in the Face of Social Inequality and Discrimination. Journal of Personality 77, 1677–1706. 10.1111/j.1467-6494.2009.00597.x

Kirmayer, L.J., Gone, J.P., Moses, J. 2014. Rethinking Historical Trauma. Transcultural Psychiatry. Volume 51, Issue 3. 10.1177/136346151453635

Knox, R., 1850. The Races of Men: A Fragment. Henry Renshaw, London.

Kohrt, B.A., Speckman, R.A., Kunz, R.D., Baldwin, J.L., Upadhaya, N., Acharya, N.R., Sharma, V.D., Nepal, M.K., Worthman, C.M., 2009. Culture in psychiatric epidemiology: Using ethnography and multiple mediator models to assess the relationship of caste with depression and anxiety in Nepal. Annals of Human Biology 36, 261–280. 10.1080/03014460902839194

Kowner, R., Demel, N. 2015. Race and racism in modern East Asia: Interactions, nationalism, gender and lineage, Chapter 1: Introduction (e-book), Brill.

Krieger, N., 2005. Stormy Weather:Race,Gene Expression, and the Science of Health Disparities. American Journal of Public Health 95, 2155–2160. 10.2105/ajph.2005.067108

Krieger, N. 2014. Discrimination and Health Inequities. Int. J. Health Services, 44 (4) 643–710.

Lee, C., 2009. “Race” and “ethnicity” in biomedical research: How do scientists construct and explain differences in health?. Social Science & Medicine 68, 1183–1190. 10.1016/j.socscimed.2008.12.036

Mannor, K.M., Malcoe, L.H., 2021. Uses of Theory in Racial Health Disparities Research: A Scoping Review and Application of Public Health Critical Race Praxis. Annals of Epidemiology 66, 56–64. 10.1016/j.annepidem.2021.11.007

Marmot, M., Allen, J.J., 2014. Social Determinants of Health Equity. American Journal of Public Health 104, S517–S519. 10.2105/ajph.2014.302200

Martin, C.L., Ghastine, L., Lodge, E.K., Dhingra, R., Ward-Caviness, C.K., 2022. Understanding Health Inequalities Through the Lens of Social Epigenetics. Annual Review of Public Health 43, 235–254. 10.1146/annurev-publhealth-052020-105613

Martinez, A., de la Rosa, R., Mujahid, M., Thakur, N., 2021. Structural racism and its pathways to asthma and atopic dermatitis. Journal of Allergy and Clinical Immunology 148, 1112– 1120. 10.1016/j.jaci.2021.09.020

McKinney, L.P., Gerbi, G.B., Caplan, L.S., Claridy, M.D., Rivers, B.M., 2020. Predictors of genetic beliefs toward cancer risk perceptions among adults in the United States: Implications for prevention or early detection. Journal of Genetic Counseling 29, 494–504. 10.1002/jgc4.1228

Mégret, F., Dutta, M., 2023. Transnational discrimination: the case of casteism and the Indian diaspora. Transnational Legal Theory 13, 391–430. 10.1080/20414005.2023.2176098

Mehra, R., Alspaugh, A., Dunn, J.T., Franck, L.S., McLemore, M.R., Keene, D.E., Kershaw, T.S., Ickovics, J.R., 2023. “‘Oh gosh, why go?’ cause they are going to look at me and not hire”: intersectional experiences of black women navigating employment during pregnancy and parenting. BMC Pregnancy and Childbirth 23. 10.1186/s12884-022-05268-9

Melton-Fant, C., 2022. Health Equity and the Dynamism of Structural Racism and Public Policy. The Milbank Quarterly 100, 628–649. 10.1111/1468-0009.12581

Narasimhan, V.M. et al., 2019. The formation of human populations in South and Central Asia. Science 365, 1–15. 10.1126/science.aat7487

Pageen Manolis Small, 2020. Achieving equity and agreement: The importance of inclusion of marginalized groups in hospital policy initiatives. Voices in bioethics 6, 1–3. 10.7916/vib.v6i.6084

Parkman, A.A., Foland, J., Anderson B., et al., 2015. Public awareness of genetic nondiscrimination laws in four states and perceived importance of life insurance protections. Journal of Genetic Counseling 24, 512–521. 10.1007/s10897-014-9771-y

Parra-Cardona, R., López-Zerón, G., Leija, S.G., et al., 2019. A Culturally Adapted Intervention for Mexican-Origin Parents of Adolescents: The Need to Overtly Address Culture and Discrimination in Evidence-Based Practice. Family Process 58, 334–352. 10.1111/famp.12381

Poliakov, L. 1974. The Aryan Myth: A History of Racist and Nationalist Ideas in Europe. Translated by E. Howard. New York: Basic Books, 1971, 1974. Pp. x+388.

Powell-Young, Y.M., Spruill, I.J., 2013. Views of Black Nurses Toward Genetic Research and Testing. Journal of Nursing Scholarship 45, 151–159. 10.1111/jnu.12015

Rasali, D.P., 2023. California’s anti-caste discrimination bill could lead to humanity’s health equity. Dignity Post, https://www.dignitypost.com/news/2023/09/99.

Rasali, D., Kao, D., Fong, D., Qiyam, L., 2019. Priority Health Equity Indicators for British Columbia: Preventable and Treatable Premature Mortality. BC Center for Disease Control, Provincial Health Services Authority, Vancouver, B.C.

Rasali, D., Li, C., Mak, S., Rose, C., Janjua, N., Patrick, D., 2021. Correlations of COVID-19 incidence with neighborhood demographic factors in BC. Annals of Epidemiology 61, 17. 10.1016/j.annepidem.2021.05.040

Reich, D., 2019. Who We Are and How We Got Here : Ancient DNA and the New Science of the Human past. Vintage Books, New York.

Relova, S., Joffres, Y., Rasali, D., Zhang, L.R., McKee, G., Janjua, N., 2022. British Columbia’s Index of Multiple Deprivation for Community Health Service Areas. Data 7, 24. 10.3390/data7020024

Sankar, P., Cho, M.K., Condit, C.M. et al., 2004. Genetic Research and Health Disparities. JAMA 291, 2985. 10.1001/jama.291.24.2985

Sims, C.M., 2010. Ethnic notions and healthy paranoias: understanding of the context of experience and interpretations of healthcare encounters among older Black women. Ethnicity & Health 15, 495–514. 10.1080/13557858.2010.491541

Soled, D.R., Chatterjee, A., Olveczky, D., Lindo, E.G., 2021. The Case for Health Reparations. Frontiers in Public Health 9. 10.3389/fpubh.2021.664783

Subica, A.M., Link, B.G., 2022. Cultural trauma as a fundamental cause of health disparities. Social Science & Medicine 292, 114574. 10.1016/j.socscimed.2021.114574

Sweeting, J.A., Akinyemi, A.A., Holman, E.A., 2023. Parental Preconception Adversity and Offspring Health in African Americans: A Systematic Review of Intergenerational Studies. Trauma, Violence, & Abuse 24, 1677–1692. 10.1177/15248380221074320

Thapa, T.B., 2014. Living with Diabetes: Lay Narratives as Idioms of Distress among the Low-Caste Dalit of Nepal. Medical Anthropology 33, 428–440. 10.1080/01459740.2012.699985

Tiedt, J.A., Brown, L.A., 2014. Allostatic Load: The Relationship Between Chronic Stress and Diabetes in Native Americans. Journal of Theory Construction & Testing 18, 22–27.

Tricco, A.C., Lillie, E., Zarin, W., O’Brien, K.K. et al., 2018. PRISMA Extension for Scoping Reviews (PRISMA-ScR): Checklist and Explanation. Annals of Internal Medicine 169, 467–473. 10.7326/m18-0850

Vervoort, D., Kimmaliardjuk, D.M., Ross, H.J., Fremes, S.E., Ouzounian, M., Mashford-Pringle, A., 2022. Access to Cardiovascular Care for Indigenous Peoples in Canada: A Rapid Review. CJC Open 4. 10.1016/j.cjco.2022.05.010

Viladrich, A., 2021. Sinophobic Stigma Going Viral: Addressing the Social Impact of COVID-19 in a Globalized World. American Journal of Public Health 111, 876–880. 10.2105/ajph.2021.306201

Wallis, M., Fleras, A., 2009. Introduction: Conceptualizing the Politics of Race: Taking Race Seriously, in: Wallis, M., Fleras, A. (Eds.), The Politics of Race in Canada. Oxford University Press, USA, pp. x–xxiv.

White, K., Lawrence, J.A., Tchangalova, N., Huang, S.J., Cummings, J.L., 2020. Socially-assigned race and health: a scoping review with global implications for population health equity. International Journal for Equity in Health 19. 10.1186/s12939-020-1137-5

Williams, D.R., Lawrence, J.A., Davis, B.A., 2019. Racism and Health: Evidence and Needed Research. Annual Review of Public Health 40, 105–125. 10.1146/annurev-publhealth-040218-043750

Yavorsky, J.E., 2019. Uneven Patterns of Inequality: An Audit Analysis of Hiring-Related Practices by Gendered and Classed Contexts. Social Forces 98, 461–492. 10.1093/sf/soy123

Yengde, S., 2021. Global Castes. Ethnic and Racial Studies 45, 340–360. 10.1080/01419870.2021.1924394

Zandy, M., Zhang, L.R., Kao, D., Rajabali, F., Turcotte, K., Zheng, A., Oakey, M., Smolina, K., Pike, I., Rasali, D., 2019. Area-based socioeconomic disparities in mortality due to unintentional injury and youth suicide in British Columbia, 2009–2013. Health Promotion and Chronic Disease Prevention in Canada 39, 35–44. 10.24095/hpcdp.39.2.01

Zhang, L.R., Rasali, D., 2015. Life expectancy ranking of Canadians among the populations in selected OECD countries and its disparities among British Columbians. Archives of Public Health 73, 1–10. 10.1186/s13690-015-0065-0

Zhou, X., Wodtke, G.T., 2019. Income Stratification among Occupational Classes in the United States. Social Forces 97, 945–972. 10.1093/sf/soy074

